# Extreme MetaboHealth scores in three cohort studies associate with plasma protein markers for inflammation and cholesterol transport

**DOI:** 10.1101/2024.12.01.24318258

**Authors:** D. Bizzarri, E.B. van den Akker, M.J.T. Reinders, R. Pool, M. Beekman, N. Lakenberg, N. Drouin, K.E. Stecker, A.J.R. Heck, E.F. Knol, J.M. Vergeer, M.A. Ikram, M. Ghanbari, A.J. van Gool, BBMRI-NL, D.I. Boomsma, P.E. Slagboom

## Abstract

The MetaboHealth score is a highly informative health indicator in ageing studies and yet contains only a small number of metabolites. Here we estimate the heritability of the score in 726 monozygotic (MZ) and 450 dizygotic (DZ) twin pairs, and test for association with plasma proteins by comparing extreme scoring individuals selected from two large population cohorts -the Leiden Longevity Study (LLS) and the Rotterdam Study (RS) and discordant monozygotic twin pairs from the Netherlands Twin Register (NTR).

The heritability for the MetaboHealth score was estimated at 40%. In 50 high and 50 low scoring MetaboHealth groups from LLS and RS, we uncovered significant differences in plasma proteins, notably in 3 (out of 15) cytokines (GDF15, IL6, and MIG), and 106 proteins (out of 289) as determined by Mass Spectrometry based proteomics analysis. A high MetaboHealth score associated with an increased level for 42 serum proteins, predominantly linked to inflammation and immune response, including CRP and HPT. A low score associated with decreased levels of 71 proteins enriched in high-density lipoprotein (HDL) remodeling and cholesterol transport pathways, featuring proteins such as APOA1, APOA2, APOA4, and TETN.

In MZ twins selected for maximal discordance within a pair we found 68 serum proteins associated with the MetaboHealth score indicating that a minor part of the associations observed in LLS and RS is likely explained by genetic influences. Taken together, our study sheds light on the intricate interplay between MetaboHealth, plasma proteins, cytokines, and genetic influences, paving the way for future investigations aimed at optimizing this mortality risk indicator.

## INTRODUCTION

As the global human population rapidly ages, it is valuable to measure vulnerability and expected resilience of older individuals to support prevention and well-informed treatment aimed at enhancing well-being [1]. Efficient disease prevention hinges on possibilities for evaluating not only an individual’s disease risk, but also the overall physiological vulnerability in an early stage which is often referred to as biological age. Originally the biological age of individuals was estimated from a suite of physiological tests and biochemical clinical quantifications [2]. More recently explorations shifted towards comprehensive molecular (‘omics’) datasets, providing global information on an individual’s biological state. Currently blood-based biomarkers to assess overall vulnerability in aging are constructed from molecular markers and based on chronological age, disease onset and mortality [3]. Here we focus on data from metabolomics and proteomics platforms representing such novel molecular markers.

Proton nuclear magnetic resonance (^1^HNMR) metabolomics enables a cost-effective and standardized assessment of a multitude of small circulating metabolites. Recent extensive collaborative efforts, like BBMRI-NL [4], FINSK/THL [5], COMETS [6], and the UK-Biobank [7], resulted in large datasets generated on the same Nightingale Health Pl ^1^H-NMR metabolomics platform. This platform has been largely explored as a source for generating markers associated with a multitude of endpoints (e.g., type 2 diabetes [8], aging [4], risk factors [9], and disease onset [10]). It gained particular attention, after training the MetaboHealth score, using mortality as endpoint, in the largest study of its kind so far (44,000 individuals and 5,500 incident deaths) [11]. This score stratifies mortality risk with a higher accuracy than conventional clinical variables, with lower and higher values indicating low and higher 5 years risk for mortality, respectively. The MetaboHealth score, though originally trained on mortality outcome, predicts multiple conditions related to overall health decline associated with ageing, such as frailty [12], cognitive decline [13], cancer, as well as respiratory deficiencies [7]. Remarkably, the MetaboHealth score includes only 14 metabolic markers. These are involved in processes like glycolysis, fatty acid metabolism, lipoproteins, and inflammation, e.g., GlycA. Although the MetaboHealth score offers an indication on physiological vulnerability, especially for older individuals, it remains largely illusive which pathophysiological mechanisms and corresponding blood factors are tracked by this mortality-trained risk score.

To address this question, we performed profiling of proteins and cytokines in serum to explore which molecular pathways co-vary with the MetaboHealth score. To this end, we tested for differences in plasma protein profiles of 50 out of 2200 Leiden Longevity Study participants (mean age∼ 56 y.o.) with the most extreme low and high values of MetaboHealth; similarly 50 out of 2900 Rotterdam Study participants (mean age∼ 67 y.o.), and 50 monozygotic twins (MZTs) from the 25 most discordant MetaboHealth scoring pairs out of 2,754 twins in the Netherlands Twin Register (mean age∼ 36 y.o.). Considering that MZTs have identical genomes, their within-pair associations are free of genetic confounding [14]. Therefore, while the first two cohorts offer an insight into population associations, the MZTs differences inform to what extent MetaboHealth differences in the plasma proteins and cytokines are unconfounded by shared genetics and environment. Overall, our exploration leads to a better understanding of the predictive power of the MetaboHealth score.

## RESULTS

### Description of the dataset and study population

We performed a nested case-control study design [15], selecting 50 participants with extreme MetaboHealth scores (MetaboHealth), 25 with high score and 25 with low score, from the middle-aged cohort Leiden-Longevity-Study Partners-Offspring (LLS_PAROFFS, mean age∼ 56 y.o.), and the Rotterdam Study (RS, mean age∼ 67 y.o.) composed by older aged individuals (Figure 1B and Supplementary Table). To minimize potential confounding, we ensured that the lower extreme samples were age- and sex-matched, with at least one high MetaboHealth case (Figure 1A-B). Given that a high MetaboHealth score corresponds to higher mortality risk and poor health status, we categorized individuals with high scores as “cases” and the remainder as “controls”. These disparities in scores are manifested also in phenotypic characteristics. We observed that the cases show significantly higher BMI in LLS_PAROFFS (cases: 26.53 vs controls: 24.37 kg/m^2^) and a higher incidence of antihypertensive medication in RS (cases: 15 vs controls: 6 users) (Supplementary Table S1). We observed in both study samples a lower level of lymphocyte percentage in the cases (RS: cases= 25.58% vs controls= 35.24%, LLS_PAROFFS: cases=24.72% vs controls= 31.52%) (Figure 1B). Interestingly, cases and controls shared similar phenotypic characteristics, despite being derived from two independent cohorts, differing for the significantly higher ages in RS (mean (age) _RS_= 74 y.o., mean (age) _LLS-PAROFFs_=59 y.o.), accompanied by a slightly elevated BMI in RS (mean (BMI) _RS_= 25.86, mean (BMI) _LLS-PAROFFs_=24.86) and most importantly the larger MetaboHealth contrast in RS (mean (MetaboHealth contrast) _RS_= 2.4, mean (MetaboHealth contrast) _LLS-PAROFFs_=1.7) (Figure 1B, Supplementary Table S1).

**Figure 1:**
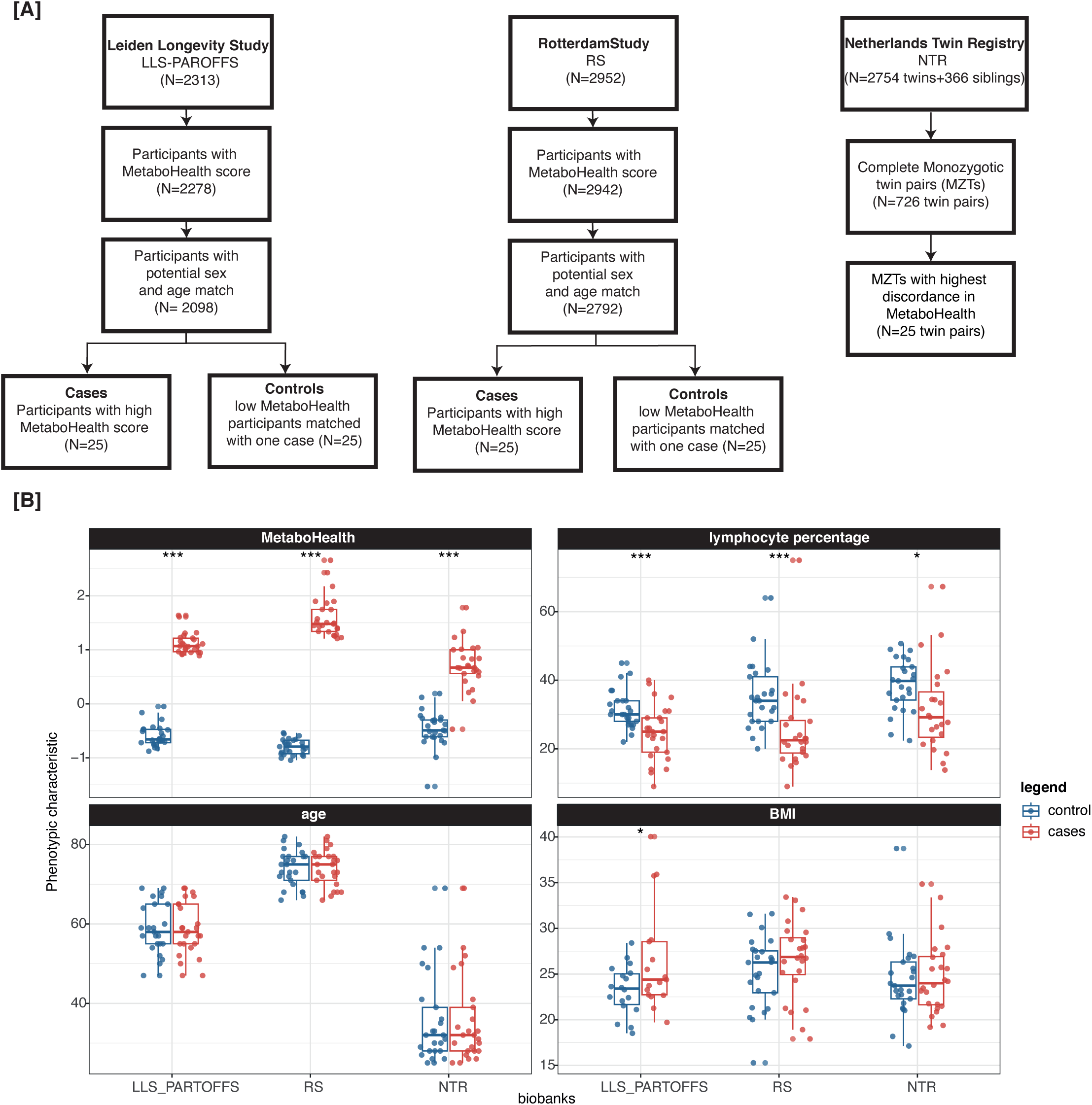
Study description. A) Flowchart detailing the inclusion criteria for samples within each study population. B) Contrast in MetaboHealth, lymphocyte percentage, age and BMI within each cohort.

To investigate to what extent associations between proteins and MetaboHealth scores reflect confounding by shared genetic or environmental factors, we investigated the contrast in MetaboHealth discordant monozygotic twins (MZTs) from the Netherland Twin Register dataset (NTR, mean age∼ 36 y.o.) (Figure 1A). 25 Twin pairs with the highest discordance with respect to their MetaboHealth score were selected from 2,754 NTR participants. In accordance with the observations in RS and LLS, the individuals with higher MetaboHealth exhibited a significant reduction in the lymphocyte percentage (cases: 31.37% vs controls: 38.64%). Nonetheless, the NTR shows some relevant characteristics when compared to the other two studies. It represents the youngest population, with a 20-year gap (mean(age)_NTR_=36 y.o.), it has a larger presence of females (36 out of 50), for which the MetaboHealth previously showed a reduced accuracy [16]. These differences, and the selection criterium based on the twin discordancy, possibly lead to a diminished contrast in MetaboHealth values (mean (MetaboHealth contrast) _NTR_= 1.18) (Figure B).

### Luminex cytokine assays: higher levels of GDF15, IL6, and MIG in the participants with a high MetaboHealth score

To explore the inflammatory state variation between the MetaboHealth extremes, we quantified 15 cytokines on the Luminex platform (Materials and Methods for a complete explanation). Six out of the 15 (40%) cytokines (IL2, TRAIL, GRO1a, IFNg, ILb, and PAI1) were below the detection threshold in most samples, likely implying that the participants were relatively healthy at the time of sampling (Figure 2A and S1). Indeed, on average, more cytokines go undetected in low MetaboHealth participants across the LLS and RS cohorts, hinting at their overall lower inflammation rate (Figure 2A). This discrepancy in detectability attains statistical significance in the case of IL6 (p-value=0.001) (Figure 2A). The same distinctive patterns between the high and low MetaboHealth score cohorts can be observed when considering the two cohorts separately (Figure S1 B-C).

**Figure 2:**
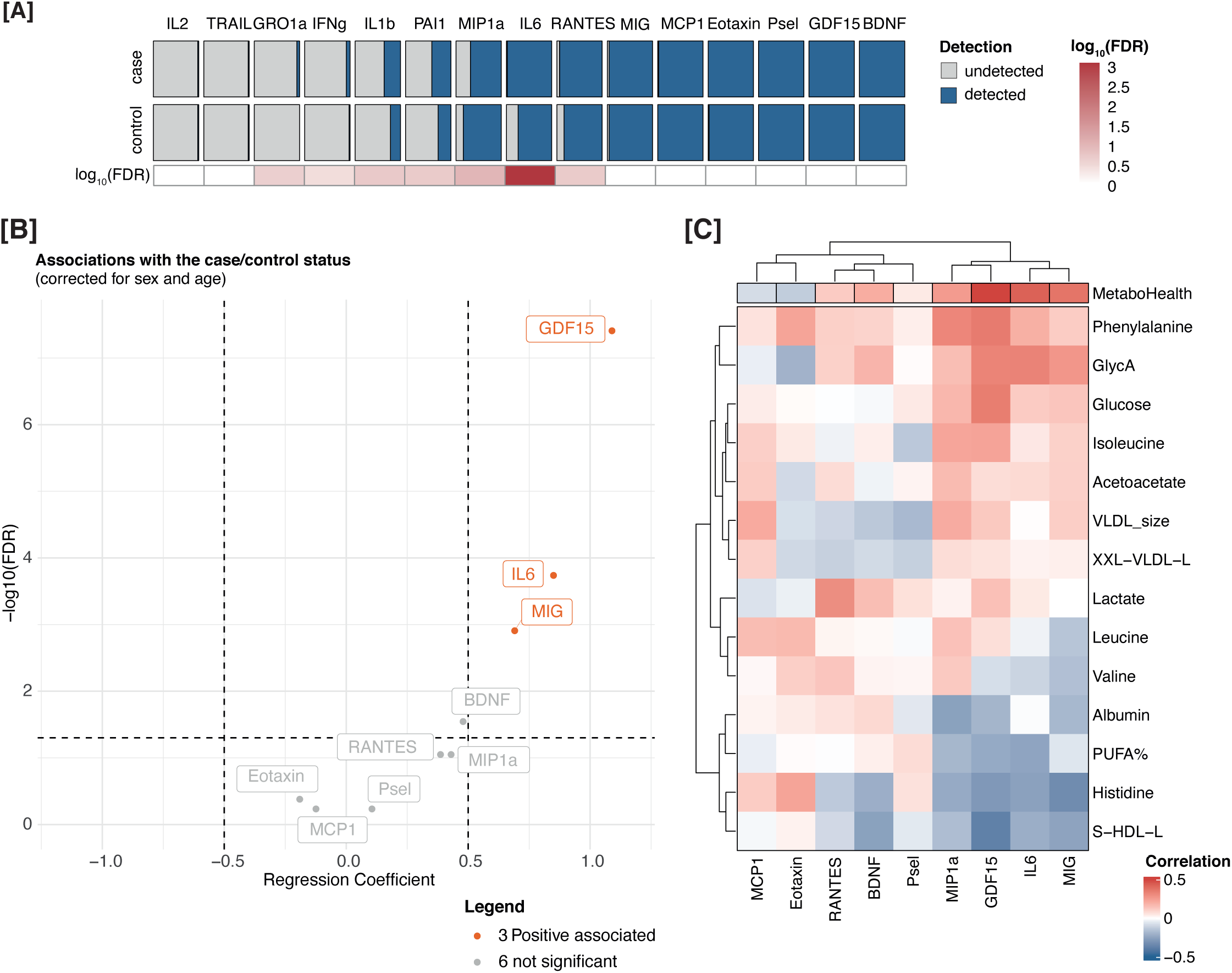
Increase GDF15, IL6, and MIG associate with MetaboHealth levels. A) Differential detectability between cases (top) and controls (bottom) for the Luminex cytokine measures, with detected values in blue and undetected in grey. The heatmap on the bottom details the adjusted p-value of the Fisher test evaluating the significances of the differential detectability. C) Volcano-plot pertaining the univariate associations between the cytokines’ levels and the participants’ case/control status corrected for sex, and age. In red the positively associated cytokines, in grey the ones not significant. D) Heatmap depicting the correlations between each cytokines with the components of the MetaboHealth score.

Our subsequent analyses focused on nine cytokines (MIP1a, IL6, RANTES, MIG, MCP1, Eotaxin, Psel, GDF15, and BDNF) exhibiting the fewest detectability issues (Figure 2A). We assessed differential expression of these cytokines between MetaboHealth cases and controls, based on a univariate linear regressions corrected for age, and sex as fixed effects. Interestingly, significantly higher levels of GDF15 (estimate∼1.08, fdr=3.9 x 10^-8^), IL6 (estimate∼1.05, fdr=1.8 x 10^-4^), and MIG (estimate∼0.95, fdr=1.23 x 10^-3^) were observed in the high MetaboHealth group when considering LLS_PAROFFS and RS together (Figure 2B). Adjusting for medication usage (blood pressure lowering and statins) and cell count (particularly lymphocyte %) influenced mostly the association with GDF15 (Figure S2A-D), yet it remained significant. Finally, reproducing the univariate associations separately for the two cohorts shows similar patterns (Figure S2F), underpinning their robustness.

To further investigate the origin of the observed signal, we looked into the correlation structure between cytokines. The generally modest intercorrelation showed a profile of mostly independent features (Figure S2E). Next, we estimated the correlation of the cytokines with the metabolomics components of the MetaboHealth score (Figure 2C). While MetaboHealth exhibits the highest correlations, we observed several noteworthy relations, i.e. strong positive correlations between Glycoprotein Acetyls (GlycA), a metabolomics inflammation marker, and GDF15 (r=0.33, p=8.2 x 10^-4^), MIG (r=0.27, p= 7.5x 10^-3^), and IL6 (r=0.33, p= 1.9x 10^-3^). Intriguingly, GDF15 displayed also elevated positive correlations with glucose (r=0.35, p= 4.2x 10^-4^), phenylalanine (r=0.35, p= 3.0 x 10^-3^), and isoleucine (r=0.23, p= 2.1x 10^-2^). In contrast, we uncovered prominent negative correlations between GDF15 and S-HDL-L (total lipids in small HDL) (r=-0.38, p= 9.9x 10^-5^), and between MIG and Histidine (r=-0.33, p= 8.7x 10^-4^).

### Plasma proteomics associated with extremes in MetaboHealth are enriched for inflammatory response and cholesterol transport pathways

In the two population-based samples (50 cases and 50 controls), we investigated plasma proteome profiles by a DIA-based quantitative plasma proteomics pipeline [17,18] (Materials and Methods). 261 out of 337 measured plasma proteins (77%) passed the detection limit and quality control criteria (detailed in Materials and Methods). We identified 106 (68 negative and 38 positive) significant univariate linear associations with the MetaboHealth extremes, adjusted for age, sex, and BMI (Figure 3A). APOA1 (estimate∼-1.46, fdr=2.44 x 10^-13^), APOA2 (estimate∼ −1.4, fdr= 4.02x 10^-12^), TETN (estimate∼-1.31, fdr= 6.42x 10^-11^), GELS (estimate∼-1.26, fdr= 3.18 x 10^-10^), and APOA4 (estimate∼ −0.93, fdr= 5.71 x 10^-6^) were the strongest negative associated proteins. In contrast, the positive acute phase proteins CRP (estimate∼1.51, fdr= 5.81x 10^-14^), LBP (estimate∼1.35, p= 1.88 x 10^-11^), HPT (estimate∼1.14, p= 1.74 x 10^-8^), were the strongest positive associated proteins. In line with what was observed for the cytokines, the additional correction for medication usage had minimal impact on the univariate associations, while lymphocyte percentage exhibited a slightly more pronounced influence (Figure S4B-D). Furthermore, a meta-analyses to evaluate the associations separately in the two cohorts revealed alike results, with generally stronger signals in the RS (Figure S5 A-B).

**Figure 3:**
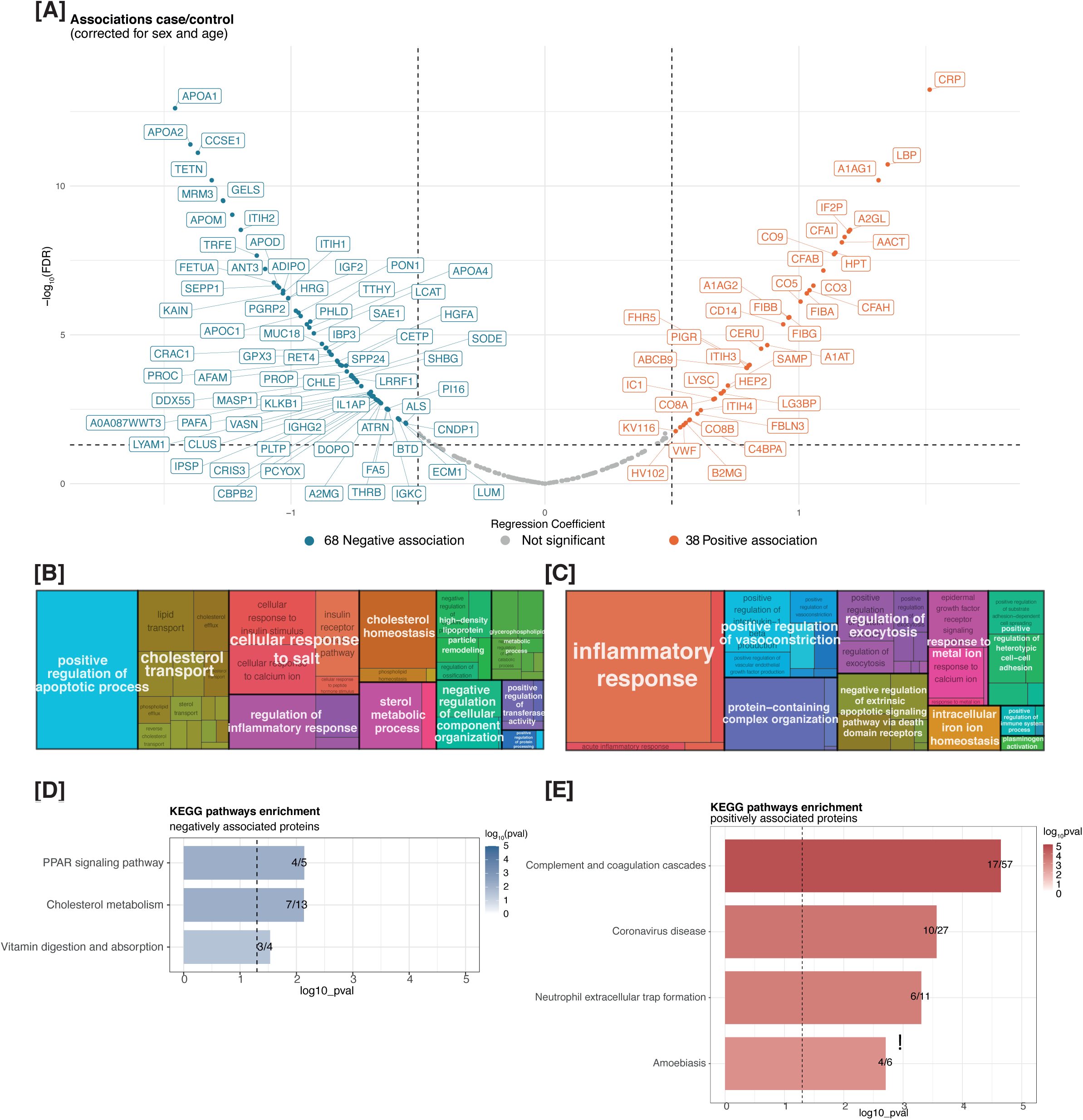
Quantitative plasma proteomics reveals proteins that associate with MetaboHealth scores in RS and LLS-PAROFFS cohorts. A) Volcano plot depicting associations of proteins with MetaboHealth scores (corrected for sex, and age). In blue the negatively and in red the positively associated plasma proteins are depicted. Grouped significantly enriched Gene Ontology Biological Processes are shown for B) the negatively and C) positively associated plasma proteins. D) and E) depict the enriched KEGG pathways for the plasma proteins respectively negatively and positively associated with the MetaboHealth score.

Subsequently, we explored the statistical interrelations and biological functionalities of the 106 plasma proteome features that exhibited significant association to the MetaboHealth extremes. These proteins showed high correlations with GlycA, comparable to those observed for the MetaboHealth score (Figure S6A). Moreover, within the group of positively associated proteins, smaller clusters of highly correlated proteins are found (Figure S6B). To gain some biological interpretation, we employed KEGG and Gene Ontology to perform functional enrichments separately for the positively and negatively associated proteins (Figure S7). As expected, considering that lower MetaboHealth values are related to healthier metabolic profiles, the negatively associated proteins demonstrated high enrichments for processes relating to “cholesterol transport”, “cholesterol metabolism”, and “high-density lipoproteins particle remodeling” (Figure 3B, S7C and S7E). Conversely, the positively associated proteins were more enriched with “Inflammatory response”, “complement and coagulation cascades”, and intriguingly, “Coronavirus disease” (Figure 3B, S7D and S7F). The latter can be interpreted as a validation, as a subset of about a dozen of inflammation related features, measured with the same proteomics platform, were previously linked to COVID19 outcome [18]. Four of these twelve COVID related plasma proteins exhibited consistently statistically significant differences between the extremes of the MetaboHealth score in RS and LLS-PAROFFs (Figure S8).

### Genetic influences on the MetaboHealth score and analysis of plasma proteins in extreme discordant MZ twins

The NTR has collected 726 complete monozygotic twin (MZTs) and 450 dizygotic twins (DZTs) twin pairs with metabolomics data. We estimated the resemblance in MetaboHealth score as a function of zygosity in these pairs. The correlation between MZ twin pairs was estimated as r= 0.432 (95% CI = 0.370-0.489), and the correlation in DZTs was r = 0.230 (95% CI = 0.141-0.316), indicating the MetaboHealth score as a heritable trait (h^2^ = 0.4) (detailed information in Materials and Methods) [19].

To exclude potential confounding from genetic factors within our associations, we conducted a monozygotic twin (MZTs) discordant twin pairs design; i.e., high and low scoring MetaboHealth genetically matched twins. In this design an observed effect is not confounded by genetic factors. Therefore, from the total population of MZ twin pairs, we selected a subsample of 25 most discordant MZ twin pairs to further explore associations of the score to the Luminex based cytokines and Mass Spectrometry-base proteomics profiling. Concordantly with the previous sections, the protein markers with the strongest associations with the MetaboHealth score also show a clear separation between cases and controls in the NTR dataset, albeit less than in the LLS and RS studies (Figure 4, and S11). To take advantage of the genetic similarity of the MZ individuals, we tested for associated proteins using a linear mixed model (Methods). The cytokines did not show significant differences in this within-pairs design, although we observed similar trends to the results in LLS and RS, with elevated cytokines in twins with high MetaboHealth scores (Figure S10A). The proteomics analyses revealed a robust signal, identifying a total of 86 significant associations (Figure S10B). Notably, CRP (estimate∼1.19, fdr= 3.75x 10^-5^) and LBP (estimate∼1.15, fdr= 8.39x 10^-5^) once again emerged as the most prominently positively associated proteins, while TETN (estimate∼-1.2, fdr= 9.69x 10^-5^) and GELS (estimate∼-1.11, fdr= 4.43x 10^-5^) were confirmed as the most negatively associated proteins. Moreover, the lower associations of APOA1 and APOA2 in the twins’ profiles differs from the contrast in LLS and RS and suggests the presence of genetic confounding in the associations between these proteins and the MetaboHealth scores. A correction for health factors had similar results as for the other two cohorts, with lymphocyte percentage having the highest effect (Figure S10C-D).

**Figure 4:**
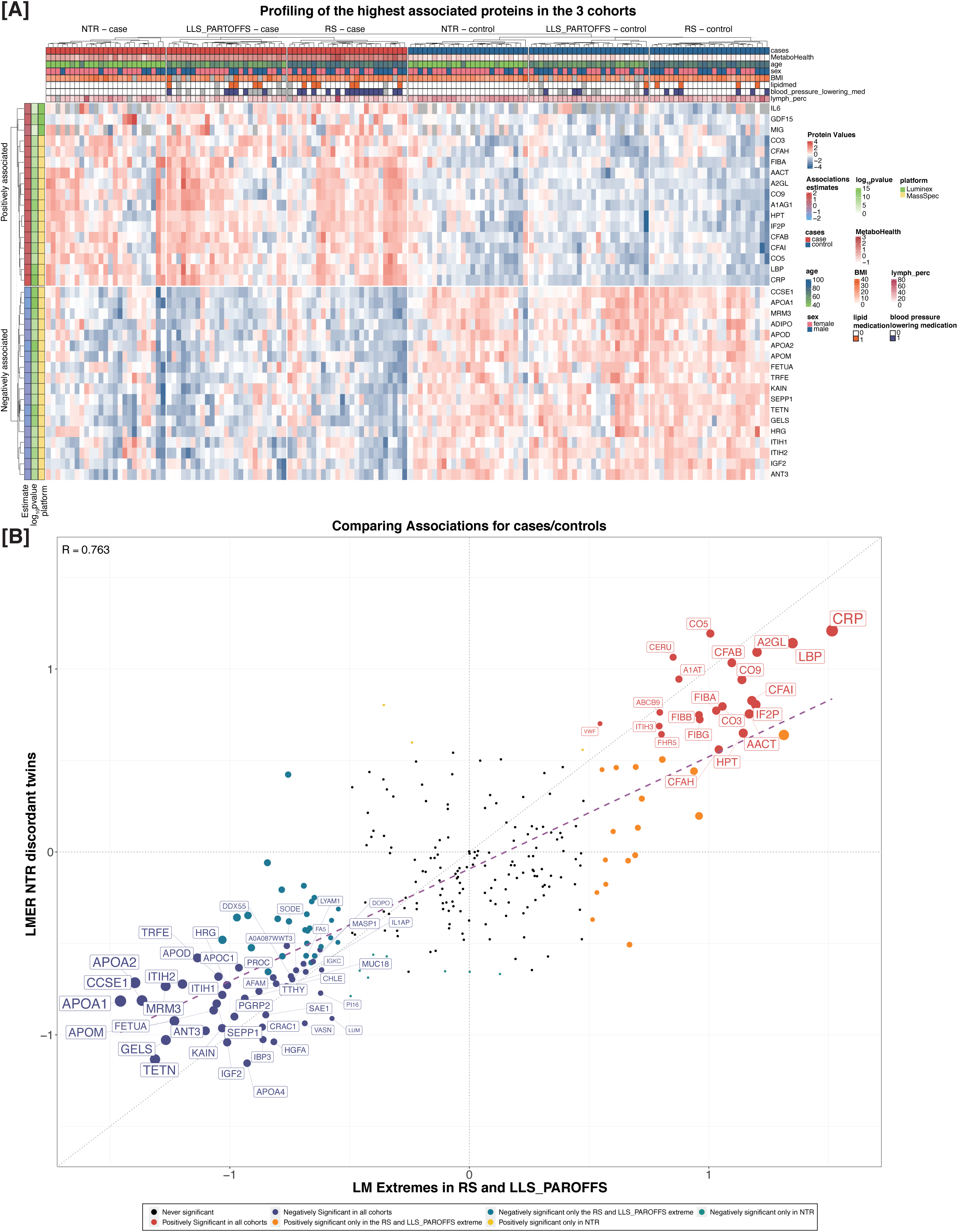
Plasma proteins associated with extreme MetaboHealth scores validated in the NTR dataset. A) Profile depicting the values of the most significantly associated proteins (|estimate|>1) and cytokines (y-axis) in all samples (x-axis), clearly separating the cases and controls in all 3 datasets. The annotation on the top show the phenotypic characteristics of all individuals. The annotation on the left shows the associations’ estimate, and log10(FDR) observed in RS and LLS, and the platform for each feature (Mass Spectrometry based Plasma Proteomics or Luminex). B) Beta-beta plot comparing the linear models in the extremes of RS and LLS_PAROFFs on the x-axis and the linear mixed models in the NTR on the y-axis. Each dots corresponds to a feature, and it’s colored based on significance. The labels are shown only when the features are consistently significant in the 2 analyses.

When comparing the NTR associations with the ones observed between the extremes in the other populations, we observed a decrease in signal but maintained a high consistency in the direction of associations (Figure 4). Specifically, 22 positively and 46 negatively associating proteins were in common with the results in RS and LLS-PAROFFs (Figure 4). These results strengthen the reliability of our previous findings indicating that the MetaboHealth score is highly informative on the overall health status of individuals, and finally that a part of its associations with protein levels in extreme individuals can be explained by genetic pleiotropy, i.e. genetic factors influencing both omics traits.

## DISCUSSION

The MetaboHealth score, along with other ^1^H-NMR metabolomics-based markers, displays risk stratification across a spectrum of health and disease outcomes relevant in ageing research. The score, though based on mortality, has shown to be an indicator of overall health status in middle and older aged individuals. Within this context, our study set out to quantify comprehensive plasma proteome profiles in 150 samples at the extreme ends of the MetaboHealth distributions from three large Dutch cohorts (Leiden Longevity Study, Rotterdam Study, and the Netherlands Twin Register, spanning a total dataset of 7,854 individuals). Our findings revealed significant differential expression among 106 plasma proteins and 3 cytokines markers, consistently observed in the RS and LLS-PAROFFs, between the highest (cases) and the lowest (controls) MetaboHealth scores, respectively indicating elevated and reduced mortality risk. These associations were for the majority not confounded by age, sex, BMI, and medication usage. A part of the proteins associated with the MetaboHealth contrast could be explained by genetic confounding as demonstrated by investigating discordant monozygotic twins.

The majority of the significant associations (68 out of 106) resulted in negatively associated proteins with the case/control contrast, indicating higher protein levels in samples of healthier subjects (reflected by lower MetaboHealth scores). Functional enrichment of these proteins revealed associations with healthy metabolism, particularly in high-density lipoprotein (HDL) remodeling and cholesterol transport pathways. Notably, the most prominent associated proteins were APOA1 and APOA2, crucial components of HDL and widely recognized as protective markers for cardiovascular disease [20,21]. It should be noted that likely a substantial genetic component drives APOA1 and 2b, since these proteins were significantly less discordant in the MetaboHealth discordant MZ twins. This aspect must be further investigated since a genetic component may be overestimated given that the MetaboHealth contrast in the (overall younger) MZ twins was rather small in comparison to the cohort studies. Interestingly, the associations with tetranectin (TETN) and gelsolin (GELS) emerged as consistently stable also in NTR. Both these proteins are under consideration as potential protective markers for various diseases, such as cancer, cardiovascular disease and neurodegeneration [22–24].

We also observed several relevant proteins that were significantly positively associated with the MetaboHealth contrast. The cytokine markers selected for this study were previously reported to strongly associate with frailty, ageing, age-related disease, and mortality [25–28]. While a large part of the cytokines could not be efficiently detected, which can be attributed to the absence of ongoing infections and acute inflammation at blood sampling, GDF15, IL6, and MIG (gene name=CXCL9), showed significant associations with higher MetaboHealth levels. These cytokines serve different roles in the body and are involved in cell signalling immune response, and inflammation. All three markers are potential biomarkers for aging-related physiological decline and frailty, where IL6 marks chronic inflammation (“inflammaging”) [29–32]; GDF-15 is a senescence associated secretory protein (SASP) and a marker of physiological stress response and mitochondrial dysfunction and MIG is the most prominent marker emerging from the inflammatory aging clock (see materials and Methods) [25]. In this regard, also the 38 significantly positively associated plasma proteins are predominantly involved in inflammatory response, complement and coagulation cascades, and COVID19. CRP, HPT and LBP, well-known biomarkers assessing the degree of inflammation and immune response, displayed the highest significant associations.

Interestingly the inflammatory component in the positively associated markers also links to COVID19 infection pathways, regardless of their infection status, considering that blood samples used in this study were drawn up to 15 years before the COVID19 pandemic. Noteworthy is that both the proteomics panel and the metabolomics assay composing the MetaboHealth score in plasma, were previously observed to be able to stratify severe cases of COVID19 [7,18]. Several plasma proteins previously related to COVID19 were significantly different also in our MetaboHealth contrast in at least one study. These include the inter- - trypsin inhibitors family, ITIH1, ITIH2, respectively positively and negatively associated, and HRG, LCAT, which are both positively associated to MetaboHealth (Figure S8A).

Significant disparities in phenotypic characteristics were observed between cases and controls across the cohorts, mostly in the higher levels of BMI and antihypertensive usage respectively in LLS and RS. However, of great importance from a health perspective seemed to be the significant reduction of lymphocyte percentage consistently among the cases in all studies, although still within healthy ranges (20-40%). Concordantly, the systematic adjustment for health and risk factors (age, sex, BMI, lymphocyte and monocyte %, lipid and antihypertensive medication usage) resulted in an attenuation of the signal to 89/106 plasma proteins and 3/3 cytokines, with the strongest effect being related to the lymphocyte percentage (Figure S4A-D). Interestingly, a decrease in lymphocyte counts and increased cytokine in blood is frequently related to a decline in immune system functions that accompanies physiological aging [33–36]. Considering the age matching within our study inclusion criteria, these observations suggest that the MetaboHealth score successfully identified individuals with accelerated biological aging given their chronological age. While the correction for cell counts partially accounted for the observed signal, it was evident that hematopoietic variation alone does not explain the observed MetaboHealth contrast in plasma proteins. Consequently, further investigation is required to elucidate whether MetaboHealth and cell count percentages can jointly be more informative in indicating health status of older individuals and secondly what the relation with these parameters and inflammation is. .

Conducting heritability analyses within the Netherlands Twin Register allowed us to establish that the MetaboHealth score has a heritability of ∼40%, at least in relatively younger ages (mean age∼36 y.o.). Therefore, the Monozygotic twin subset provided an ideal setting to further examine the proteomics associations within a genetically identical population. Cytokines did not exhibit significant associations to the MetaboHealth contrast in the twins, while we found up to 46 negative and 22 positive plasma protein relations. An attenuation of the number and strength of associations indicate that part of the signal may be explained by genetic factors that influence both the MetaboHealth score and quantified proteins. Further research should be focused on investigating the genetic confounding in twins of higher ages where MetaboHealth is even more informative. Pertaining to this, it is essential to highlight that NTR participants had lower MetaboHealth contrasts as they are younger of age as well as due to the inclusion restriction. Supposedly for the same reasons, diminished associations are noted also when comparing the results in LLS-PAROFFs to RS (Figures S5). Possibly as individuals age, environmental and/or stochastic factors gain greater importance on the features included in the MetaboHealth score over genetic influences.

One aspect of our study that can be regarded as a limitation is the generally healthy state of the participants in the cohorts from which we derived our participants. Consequently, the observed contrast between the extreme levels of MetaboHealth was relatively modest. This is underscored by the fact that several of the examined cytokines consistently fell under the detection limit, a circumstance that can be largely attributed to the overall absence of ongoing infections at blood sampling which proteins may be useful biomarkers in clinical studies. We envision MetaboHealth as a score potentially giving an indication of vulnerability in individuals from the population in a modifiable health phase, way before onset and diagnosis of specific diseases. The score indeed effectively identified relevant biological differences within all three populations indicating that early changes in multiple metabolic and proteome parameters known to reflect decline in health is represented by the score. Given the limited number of individuals in our selection we do not have the statistical power to explore concrete endpoints such as mortality or frailty in this study. We consider this study as a proof of principle design to explore the relevance of omics scores generated in the ageing field in the context of additional omics levels, to better understand why a score predicts endpoints and by what parameters the predictive power may potentially be improved.

In conclusion, our study confirms MetaboHealth as a robust marker for the inflammatory aspects of aging. In accordance with the current biological aging theories, this score effectively identifies individuals exhibiting reduced lymphocyte counts and increased levels of pro-inflammatory proteins regardless of chronological aging. We believe that this investigation supports integrating the MetaboHealth score with proteome data to enhance the prognostic value of the score, increase our comprehension of the aging process and loss of health in individuals identified by the score and the health gain that may be expected by timely intervention.

## MATERIALS AND METHODS

### Study design

This study was actuated starting from the metabolomics data included in the BBMRI-NL consortium originated from the participants to three cohorts: Leiden Longevity Study (LLS-PAROFFS), Rotterdam Study (RS), and the Netherlands Twin Register (NTR). The Leiden Longevity study is a population-based cohort with a unique two-generation design, examining 421 Dutch long-lived families [37]. The current work was performed on the first measurements (IOP1) of the second generation, namely the Offspring and their Partners, for a total of 2,313 participants. The Rotterdam Study is a population-based prospective study on individuals living in a specific neighborhood in Rotterdam, prone to cardiovascular endpoints [38]. The current study was set off utilizing the first measurements (RS-I), which enclosed a total of 2,986 participants. The Netherlands Twin Register is a prospective study investigating young and adult twins and multiples along with their family members. In this particular study we focused on the monozygotic dizygotic twin pairs within in the cohort, for heritability estimation and implemented a within pairs case-control study design for the 25 most discordant MZ twin pairs.

### Data and sample collection

#### Metabolomics measurements

The metabolomics dataset was generated by the BBMRI-NL Consortium on the EDTA plasma samples of the entire cohorts (LLS-PAROFFS: 2,313 samples, RS: 2,986 samples, and NTR: samples). The features were quantified using the high-throughput proton Nuclear Magnetic Resonance (^1^H-NMR) platform made available by Nightingale Health Ltd., Helsinki, Finland. This technique can quantify over 230 metabolic features in a single assay, including lipids, lipoproteins, fatty acid composition, various amino acids, and their derived measures (e.g., ratios) [39,40]. We employed the dataset quantified in 2014 to ensure the correct projection of the MetaboHealth model, originally trained on this version of the platform.

### Selection of the participants from the large population studies

#### LLS-PAROFFS and RS

The sample selection was based on the chronological age independent part of the MetaboHealth, which was obtained as the residual from a linear regression of chronological age on the metabolomics score. The cases were defined as the 25 participants with the highest MetaboHealth, indicating an unhealthier status, within each cohort separately. Following, to limit the confounding effect of sex and age, the controls were selected as the participants with the lowest score that could have at least one match with the cases in terms of both age, and sex.

#### NTR

The twin population of NTR was composed on 726 complete monozygotic (MZTs) and 450 complete dizygotic (DZTs) twin pairs. We extracted the 25 twin pairs which showed the largest differences in MetaboHealth score.

### Cytokines quantification

We used previously validated multiplex immunoassays (Luminex platform) to determine plasma protein levels [41]. All assays were performed at the ISO-certified multiplex core facility of the UMC Utrecht, Utrecht, The Netherlands. Before analysis all samples were centrifuged through 0.22 μm spin-X filtration columns (Corning, Corning NY USA) to remove debris. Non-specific (heterophillic) antibodies, which may interfere with the assay, were blocked using Heteroblock (Omega Biologicals, Bozeman, MT, USA) as previously described [10,11]. If applicable, samples were diluted in high performance elisa buffer (HPE buffer, Sanquin, Amsterdam, the Netherlands). We determined levels of the immunoregulating proteins IL-1β, IL-2, IL-6, IFN-γ, GDF-15, CCL2/MCP-1, CCL3/MIP-1α, CCL5/RANTES, CCL11/Eotaxin, CXCL1/GRO-1α, CXCL9/MIG, PAI-1, BDNF, TRAIN and soluble P-selectin in plasma. These markers were selected based on two critera, first that they could be measured on our budgetary boundaries using the Luminex platform in the Utrecht University; secondly because they were previously indicated as markers of ageing, mortality, frailty and age-related disease. The last criterium was in great part based on the following studies: A) belonging to the top 15 most informative variables in the iAge clock [25](for the cytokines: CXCL9/MIG, EOTAXIN, CCL3/Mip-1α, IL-1β, IFN-γ, CXCL5/RANTES;,CXCL1/GRO-1α, CCL2/MCP1, IL-2; TRAIL, PAI-1); B) recognized as indicators of frailty (Pselectin and BNDF) [26]; C) or marker of chronic inflammation (IL6) [27], and d) finally, widely explored marker for aging, cancer, cardiovascular, and lung disease (GDF-15/MIC-1) [28].

Although the majority of the features (73%) were quantified in undiluted samples, PAI1 was measured with a dilution rate of 1/10, and, GDF15, RANTES, and BDNF, with a dilution rate of 1/100.

### Protein digestion

Protein digestion was performed on an Agilent Bravo liquid handling Platform following Vollmy et. al. [18]. A pooled QC sample was prepared by pooling an equal amount of each digested sample. Then all samples were diluted 40 times with TFA 1% to bring them to an approximate concentration of 10 ng/uL. Finally, the samples were loaded onto Evotips (Odense, Danemark) using an Agilent Bravo liquid handling platform.

### LC-MS data acquisition

The digested samples were measured using the 60 SPD method of a Evosep One (Odense, Danemark) on a EV-1109 column (C18, 8 cm x 150 µm, 1.5 µm, Evosep, Danemark) coupled to a timsTOF-HT (Bruker, Germany) equipped with a Captive Spray source and operating in DIA-PASEF adapted from Skowronek et. al. [42] Briefly, two ion mobility windows per dia-PASEF scan with 12 variable isolation window widths adjusted to the precursor densities were used. The ion mobility range was set to 0.6 and 1.5 cm^−1^. The accumulation and ramp times were specified as 100 ms for all experiments. Source capillary voltage and temperature were set to 1800 V and 180°C. Drying and sheath gas were set to 3 L/min. The pooled QC samples were injected every 8 samples.

### Processing of proteomics data

Raw data were processed using DIA-NN 1.8.1 [43], Peptides were searched against an in-silico predicted library computed from the human proteome with isoforms (UniProtKB and TrEMBL, 103830 protein entries and 20560 genes) and the common protein contaminants with 2 missed-cleavages and no variable modification. Match Between Run was used and the Heuristic inference was disabled. MS1 and MSMS mass accuracy were set to 10 and 20 ppm respectively.

The protein intensities were computed using the maxLFQ algorithm implemented in the DIA-NN R-package. For this only the prototypic precursors that satisfy the following criteria were considered: Q.value ≤ 1%, missing value ≤ 20% and RSD ≤ 40% in the QC samples. The protein groups were filtered at lib.Q.Value ≤ 1%, lib.PG.Q.value≤ 1%.

### Covariates

Data on age (in years), sex (males/females), BMI (kg/m^2^), cell counts (%), lipid medication, and blood pressure lowering medication were reported within the BBMRI-nl Consortium. These covariates were evaluated as possible confounders as they are known to be associated with both the metabolomics and the proteomics datasets. Age and sex were self-reported, and BMI was calculated based on weight and height. The cell counts percentage was defined as the measured monocytes and lymphocytes percentage, taking the granulocyte percentage as given.

## Statistical Analyses

### Preprocessing

#### Metabolomics and MetaboHealth score projection

We applied the same quality control previously described by Deelen et al. [11], using the R package MiMIR [44]. While the Nightingale Health platform measures over 250 metabolomics features, we focused our attention on the 14 metabolomics variables included in the MetaboHealth model (list of analytes can be found in the Supplementary Table S1). Then, we applied a logarithm transformation to the analytes, while adding a value of 1 to all analytes containing any zero as a value. Afterwards a z-scale normalization was applied separately within each cohort to minimize batch effects. Finally, we projected the MetaboHealth score using the coefficients indicated by Deelen et al. [11,44].

#### Cytokines

Initially, we assessed the occurrence of samples reported as under the lower detection thresholds, comparing cases and controls, separately for each feature. Subsequently, we employed the Fisher test to determine the statistical significance of these difference (p-value 0.05). Following, we proceeded to exclude six out of fifteen features that consistently fell below the lower detection thresholds in most of the quantified samples (namely, IL2, TRAIL, GRO1a, IFNg, IL1b, and PAI1 were undetected in more than 65% of the samples). There were no values recognized as outlier samples, considered as values more than 5 standard deviations (SD) away from the mean of the feature. Finally, the remaining nine features were first log transformed and then z scaled separately per biobank to reduce batch effects.

#### Proteomics

We applied a similar approach to the proteomics. We evaluated the differential patterns in missing values of the features between the cases and controls. Features with more than 5% missing values (20 missing) were subsequently excluded, resulting in a set of 261 analytes, from the 337 initial set 8 values (0.03%) were recognized as outliers, meaning that they resulted as the values 5 SD away from their mean of the feature and set as missing information. We imputed the remaining 114 missing values (0.4%) using the non-linear iterative partial least squares method (nipals), implemented in the R package *pcaMethods*. Finally, to enhance comparability and facilitate downstream analysis we performed a log transformation and a z-scaling of the features per biobank.

### Linear Association and Meta-analyses in RS and LLS-PAROFFS

Initially, we applied linear regression models across the entire dataset to assess the associations between each cytokines and proteomics features separately, with the case/control status of the participants. These analyses accounted for potential confounding factors (age, sex, BMI, cell counts, and usage of lipid medication and blood pressure lowering medication). Subsequently, we attempted to evaluate the associations independently for each cohort (LLS-PAROFFS and RS) and conducted a meta-analysis. The meta-analysis was performed with a restricted maximum likelihood estimator using the package *metafor* in R. To correct for multiple testing, we applied False Discovery Rate (FDR).

### Enrichment Analyses and Network analysis

We performed the functional enrichment of the most interesting proteomics features using the web-tool *Enrichr* [45]. We evaluated the Gene Ontology (GO) Biological Processes (BP), the KEGG pathways, and Reactome. Firstly, we performed an enrichment analyses on the full set of 337 proteomics features to evaluate the functions overall characterizing to the proteomics platform (Figure S7A-B). Secondly, we analyzed separately the 38 positively and the 68 negatively associated proteins to the MetaboHealth’s extremes corrected for sex, age, and BMI (Figure S7C-H). To ensure a fair enrichment analysis of the significantly associated proteins we used the full list of 337 proteins as the background of possible analytes. To better interpret at the GO BP results we used the R package *rrvgo* (threshold=0.7), which is able to summarize the redundant information in the database [46].

### Analyses in the Netherlands Twin Register

First, we examined heritability of the MetaboHealth score within Netherland Twin Register. Monozygotic twins (MZTs) share their (almost) complete DNA sequence, while dizygotic twins (DZTs) share on average 50% of their segregating genes. Any differences in correlations between MZTs and between DZTs offers a first hint on genetic influences in the signal. We obtained an estimate of the heritability of a trait (h^2^) using the formula: h^2^ = 2(r_MZTs_-r_DZTs_), where r denotes the correlation between the twins [19].

Secondly, we assessed the associations between the cytokines and serum protein features with the case/control status of the selected MZTs with the highest MetaboHealth differences. To do this we used linear mixed models to consider the family status, while allowing to systematically correct for potential covariates. Finally, FDR to correct for multiple testing.

### Data sharing

Mass Spectrometry data have been deposited to the ProteomeXchange Consortium via the PRIDE partner repository with the dataset identifier PXD057946. Phenotypic information are available upon reasonable request at BBMRI-nl https://www.bbmri.nl/services/samples-images-data.

## Data Availability

Mass Spectrometry data have been deposited to the ProteomeXchange Consortium via the PRIDE partner repository with the dataset identifier PXD057946. Phenotypic information are available upon reasonable request at BBMRI-nl.

https://www.bbmri.nl/services/samples-images-data

## Acknowledgements

This work was performed within the BBMRI Metabolomics Consortium funded by: BBMRI-NL (financed by NWO 184.021.007 and 184.033.111), X-omics (NWO 184.034.019), VOILA (ZonMW 457001001) and Medical Delta (METABODELTA: Metabolomics for clinical advances in the Medical Delta). EvdA is funded by a personal grant of the Dutch Research Council (NWO; VENI:09150161810095). Acknowledgements for all contributing studies can be found in the Supplementary Material-BIOS Consortium. Finally, we would like to thank Prof. Peter Bram ‘t Hoen for critically appraising this manuscript.

## Contributors

PES, DB, EbvDA, and MJTR conceived and wrote the manuscript. DB performed the analyses. EBvDA and MJTR verified and supervised the analyses. PES, MB, DiB, AjvG, DB, EbvDA, and MJTR were involved in defining the study design. PES, MB, and NL were involved in the data acquisition for the LLS-PAROFF cohort; RP and DiB in the data acquisition of the Netherland Twin Register; JmV and MG in the data acquisition of the Rotterdam Study. AjrH, ND, and KeS performed the Mass Spectrometry proteomics. EFK performed the cytokine quantification. All authors discussed the results and contributed to the final manuscript.

## Supplementary Figures legends

**Figure S1: Quality control of the cytokines in LLS-PAROFFS and RS.** A) Percentage of undetected values for each cytokine (x-axis) divided per cohort (y-axis). Differential detectability analysis performed separately for B) LLS-PAROFFS and C) RS. Cytokines distributions C) before and D) after the pre-processing steps (log transform and z-scaling).

**Figure S2: Sensitivity analyses and Meta-analyses of the cytokines’ associations with MetaboHealth extremes:** A-D) Volcano-plot of the univariate linear associations between the cytokines and the case/control status of the participants systematically corrected by increasing covariates. In grey the not significant cytokines and red the positively significant ones. E) Intercorrelations of all the cytokines. F) Forest plots of the univariate linear associations divided in RS (green), LLS-PAROFFS (yellow), and Meta-analyses (red).

**Figure S3: Quality control of the proteomics dataset in RS and LLS-PAROFFS:** A) Percentage of missingness in the highly missing features divided per cohort. B) Differential detectability between cases (top) and controls (bottom) for the proteins showing significant difference. The heatmap on the bottom details the adjusted p-value of the Fisher test evaluating the significance of the differential detectability. 9 randomly selected plasma proteins distributions C) before and D) after the pre-processing steps (log transform and z-scaling).

**Figure S4: Sensitivity analyses and Meta-analyses of the plasma proteins with the MetaboHealth extremes in RS and LLS-PAROFFS:** A-D) Volcano-plot of the univariate linear associations between the cytokines and the case/control status of the participants systematically corrected by increasing covariates. In grey the not significant cytokines, in blue the negatively and red the positively significant plasma proteins. Upset plot detailing the amount of significantly D) negatively and E) positively associated features after correcting for covariates.

**Figure S5: Meta-analyses of the associations between the proteomics features and the MetaboHealth extremes:** Forest-plot of the meta-analysis for the significantly A) Negatively and B) positively associated features.

**Figure S6: Correlations of the significantly associated proteomics:** Heatmaps of the correlations of the proteomics features with A) MetaboHealth and its components, and B) with the proteomics features themselves.

**Figure S7: Functional Enrichments of the proteomics datasets:** Bar-plots displaying the log10pvalues of the significant enrichments performed for all 320 features in the platform in A) GO Biological Processes pathways, and B) KEGG pathways. GO Biological Processes significantly enriched in the C) negatively and D) positively associated proteomics features. Reactome pathways significantly enriched in the E) negatively and F) positively associated proteomics features.

**Figure S8: Covid related proteins.** Plots displaying the case/control (red/blue) differences in RS and LLS-PAROFFs for the proteins previously associated to covid infection by Völmy et al.

**Figure S9: Pre-processing in NTR.** A) Percentage of undetected values per cytokines (y-axis) per sample (x-axis). B) Percentage of undetected values per proteomics feature (y-axis) in each sample (x-axis). C) Differential detectability of the cytokines in NTR.

**Figure S10: Associations in NTR.** Volcano plot of the univariate linear mixed models comparing the case/control status with A) cytokines and B) proteomics features corrected for age, and sex. Upset plots of the significantly C) negative and D) positive proteomics markers.

**Figure S11: Complete Heatmap profiles of the significant proteins.** Profile depicting the values of the significantly associated proteins (FDR<0.05) and cytokines (y-axis) in all samples (x-axis), clearly separating the cases and controls in all 3 datasets. The annotations on the top show the phenotypic characteristics of all individuals. The annotation on the left shows the associations’ estimate, and log10(FDR) observed in RS and LLS, and the platform for each feature (Mass Spectrometry or Luminex).

