## Supplementary Figures for "Extreme MetaboHealth scores in three cohort studies associate with plasma protein markers for inflammation and cholesterol transport"

**Figure S1:** Quality control of the cytokines

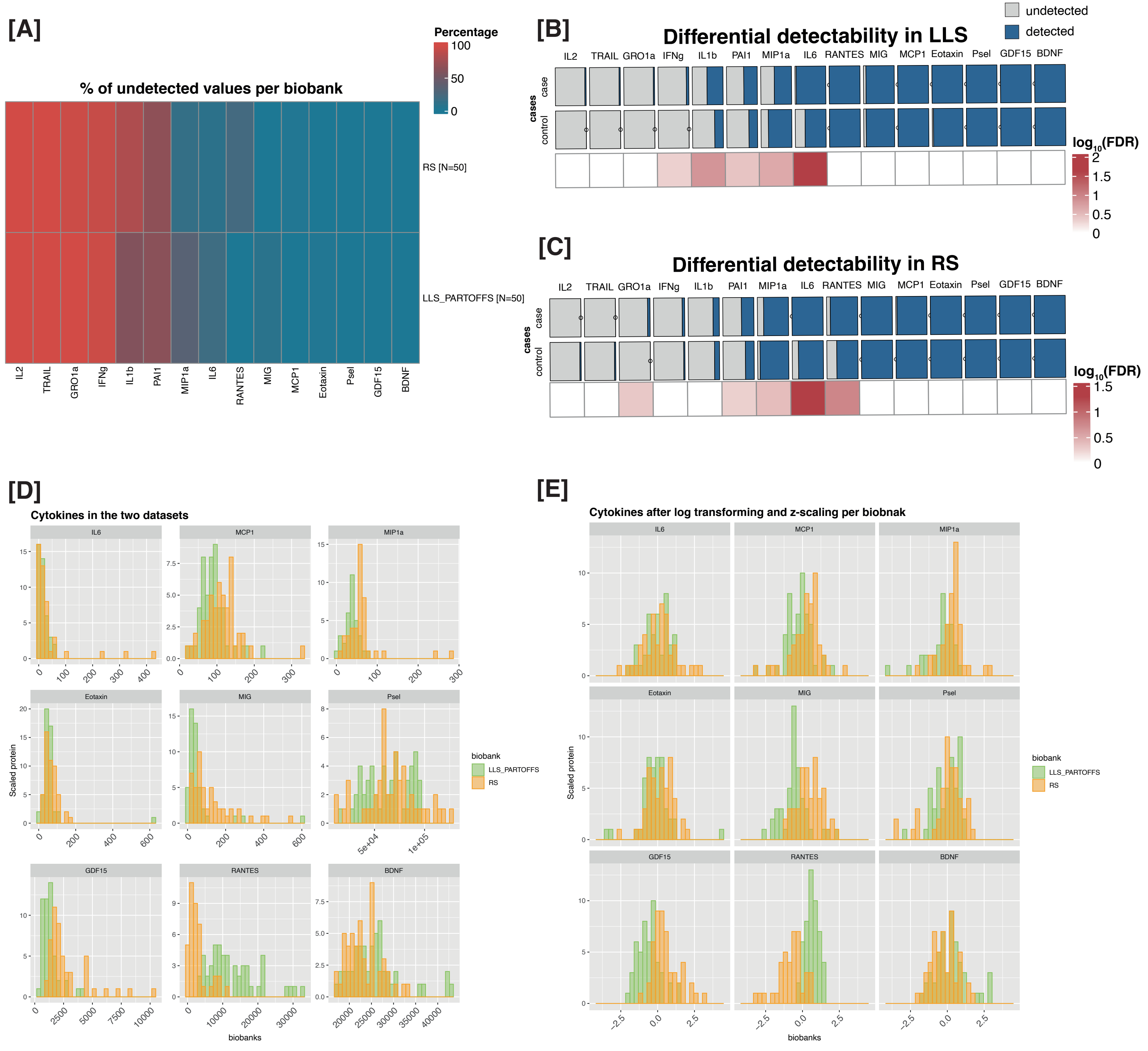

**Figure S2:** Sensitivity analysis and Meta-analysis of the cytokine's associations with the MetaboHealth extremes

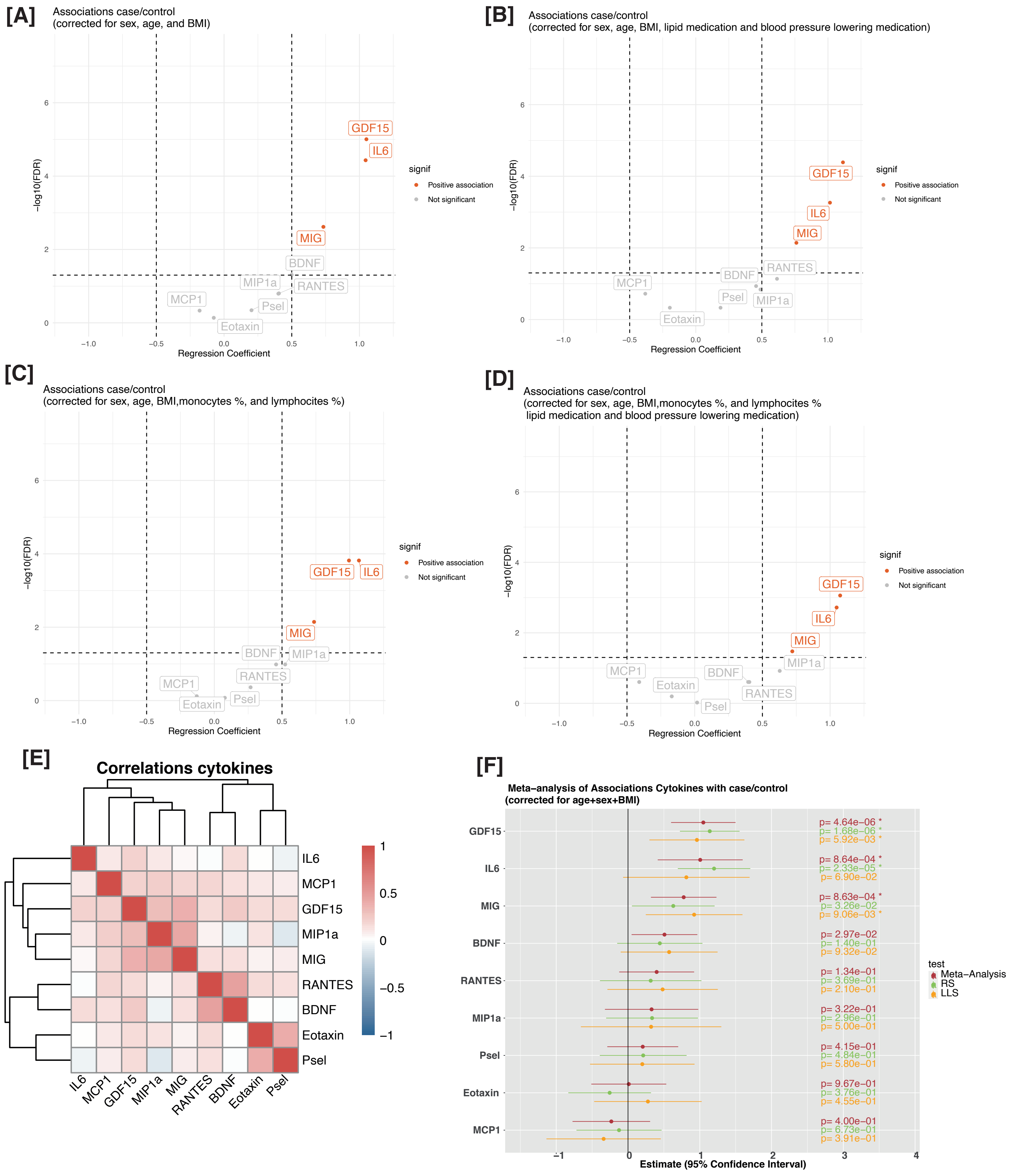

**Figure S3:** Quality control of the proteomics dataset

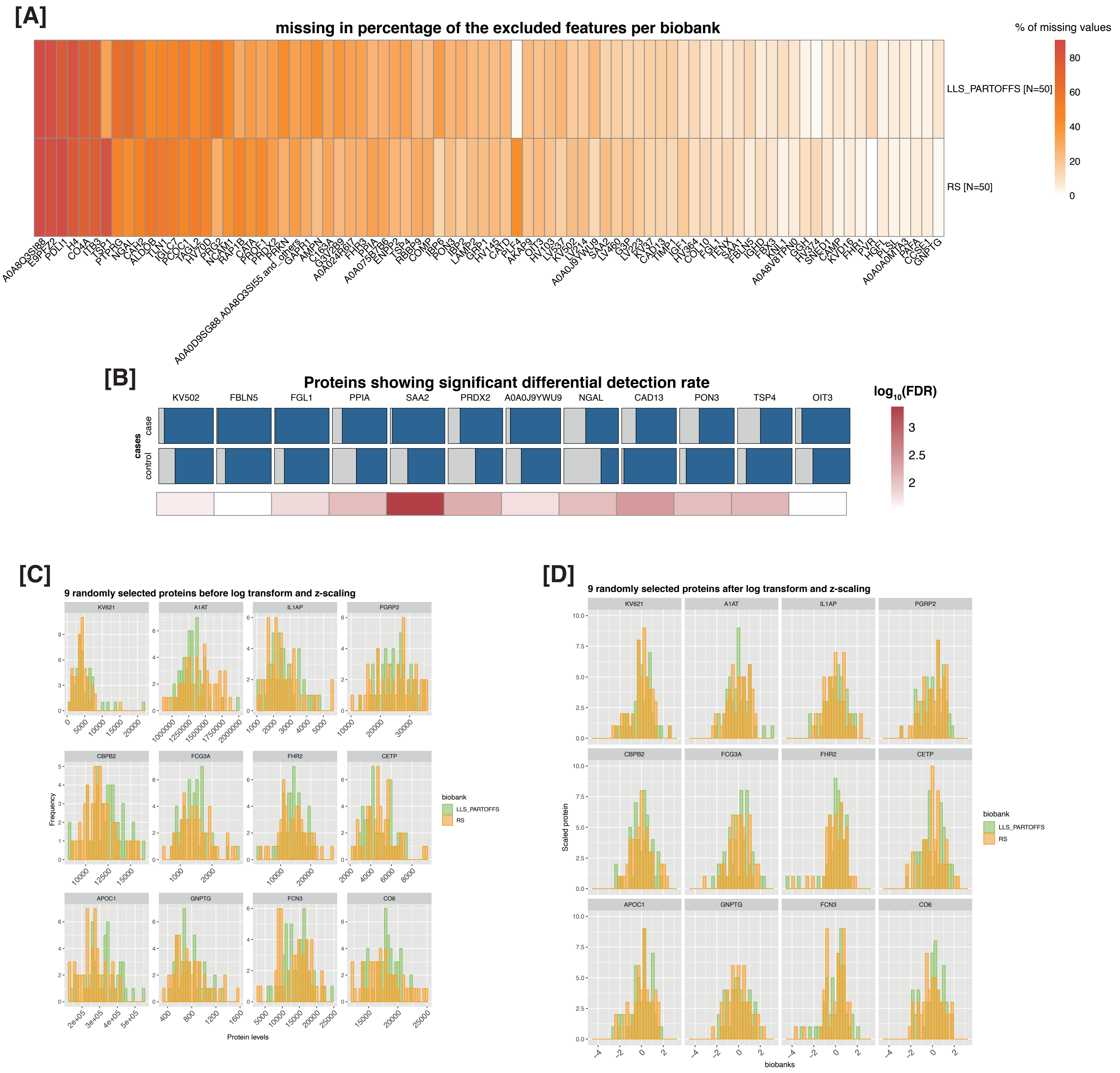

**Figure S4:** Sensitivity analyses and Meta-analyses of the plasma protein with MetaboHealth extremes in RS and LLS-PAROFFS

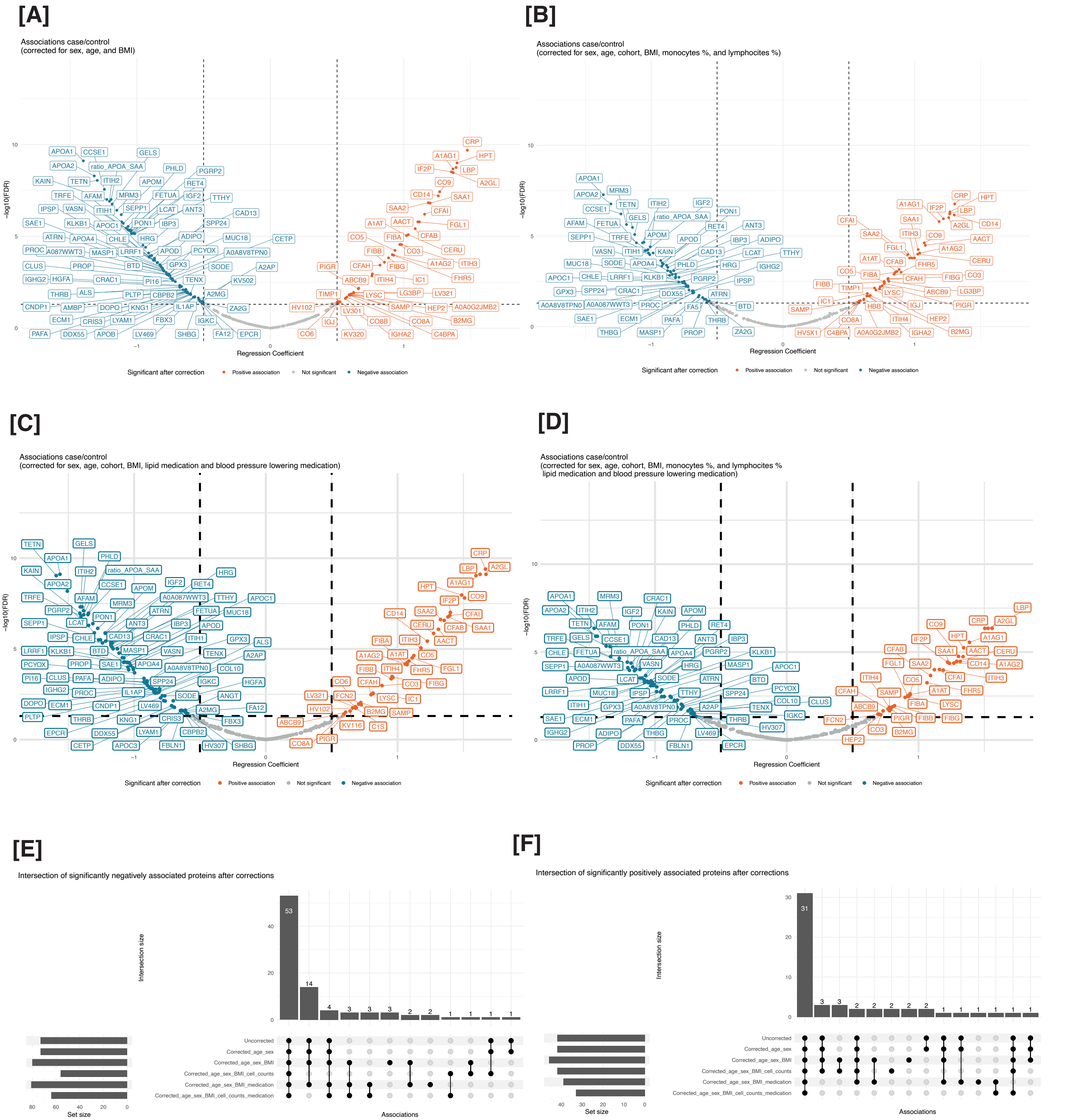

**Figure S5:** Meta-analyses of the associations between the proteomics features and the MetaboHealth's extremes

[A]

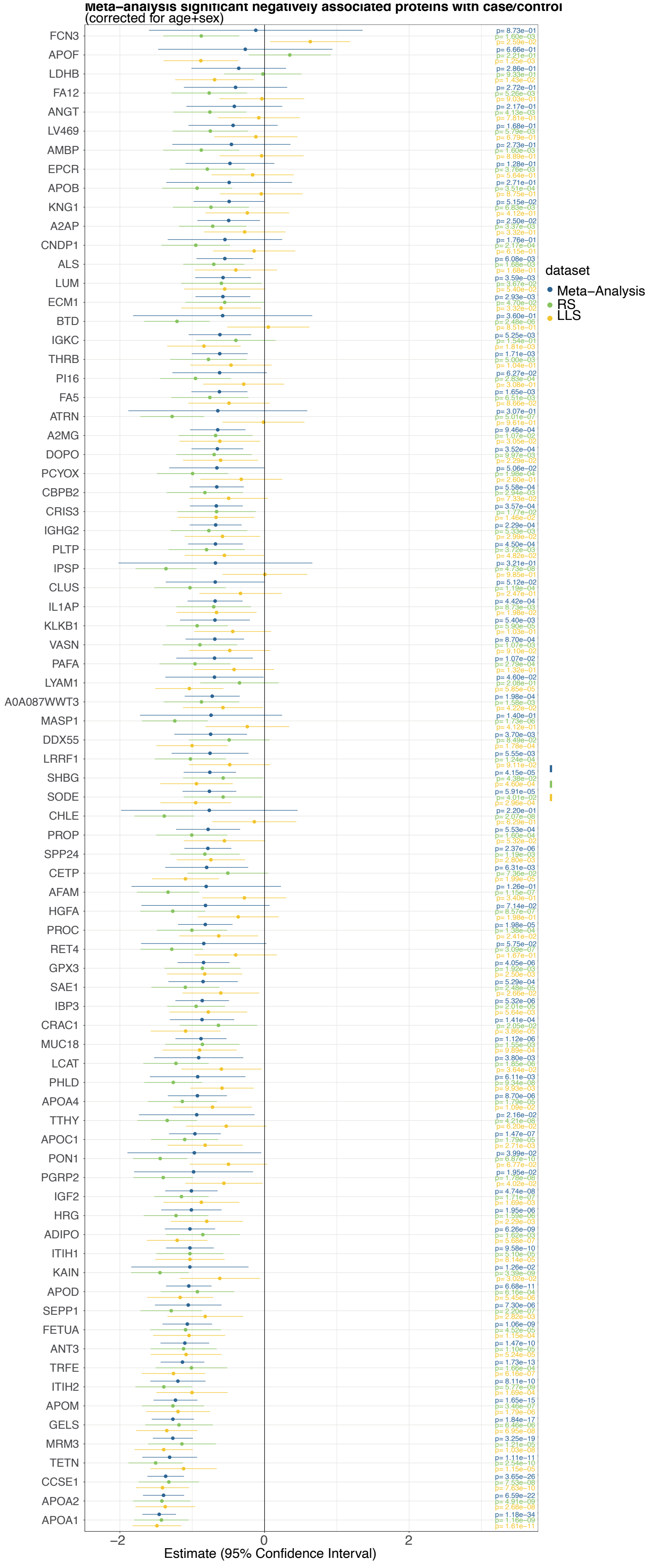


[B]

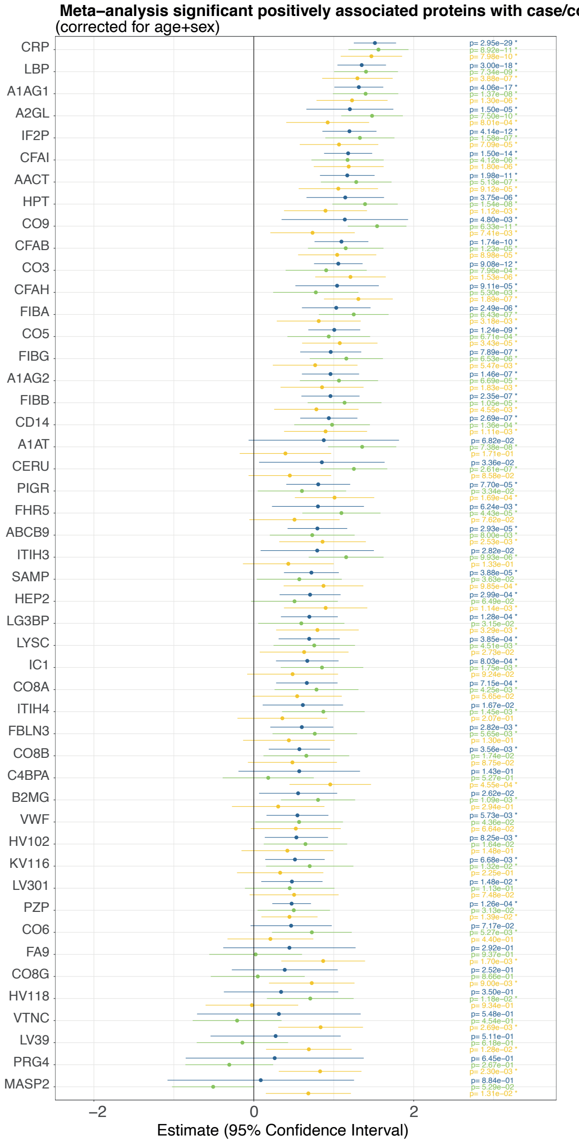

**Figure S6:** Correlations of the significantly associated proteins

**[A]** **[B]**

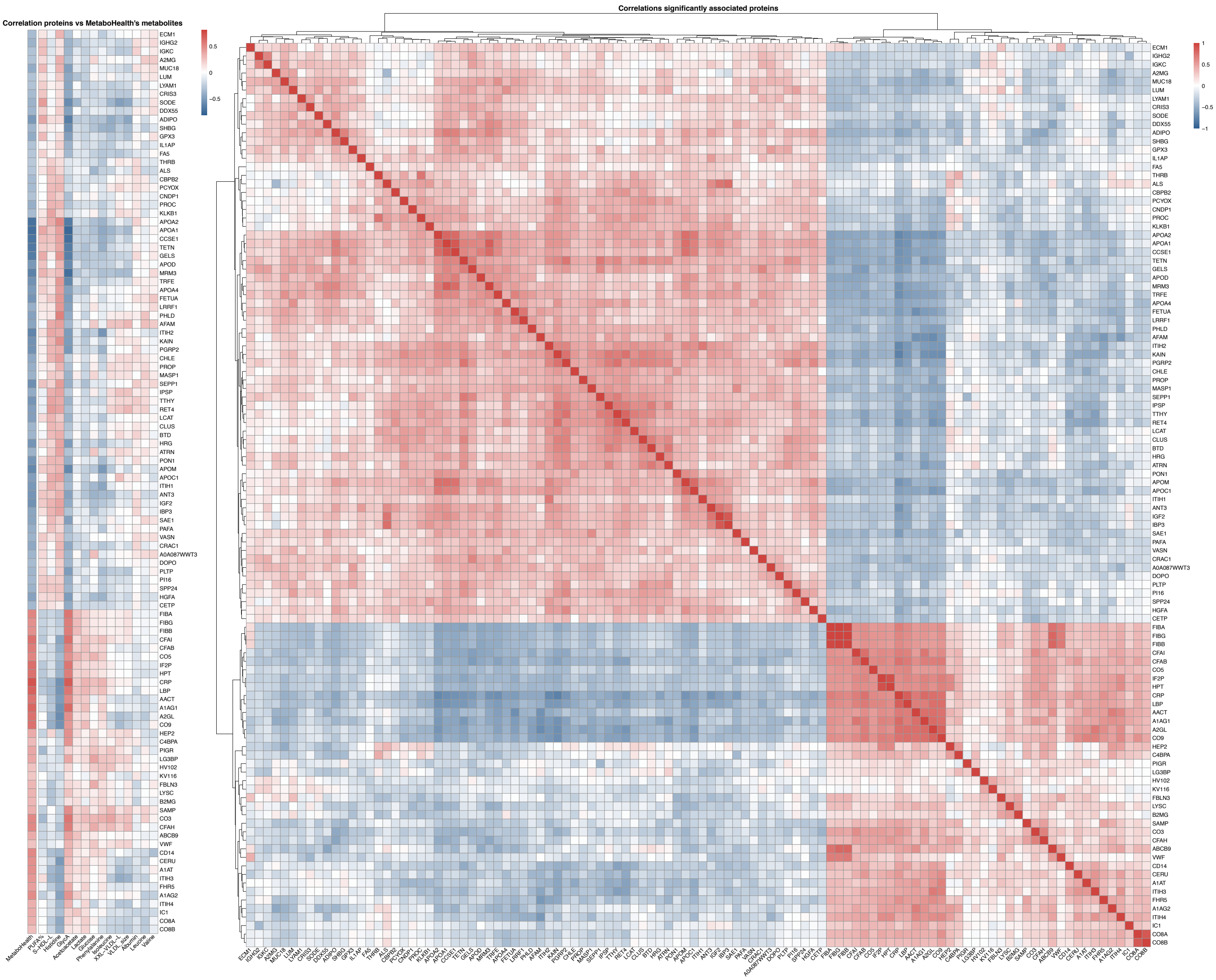

**Figure S7:** Functional enrichments in the proteomics dataset

[A]

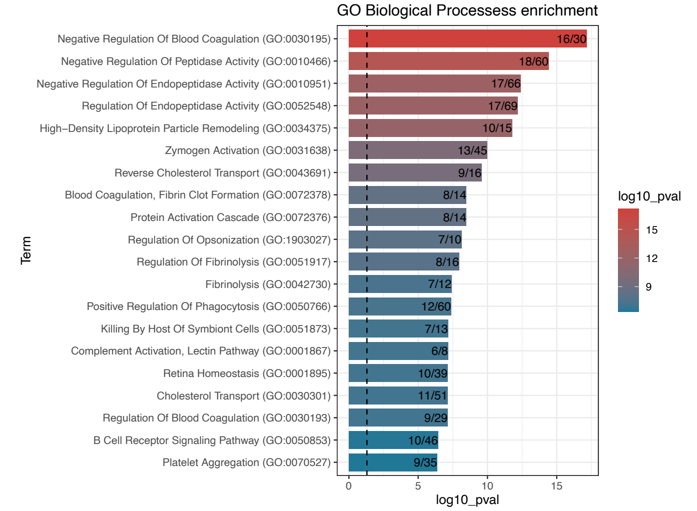


[B]

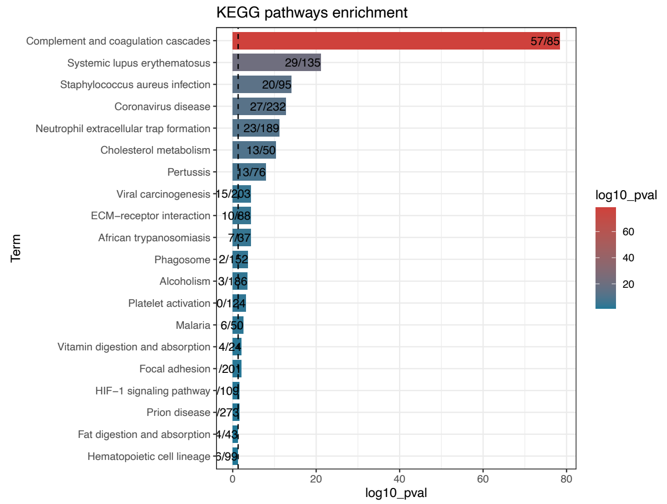


[C]

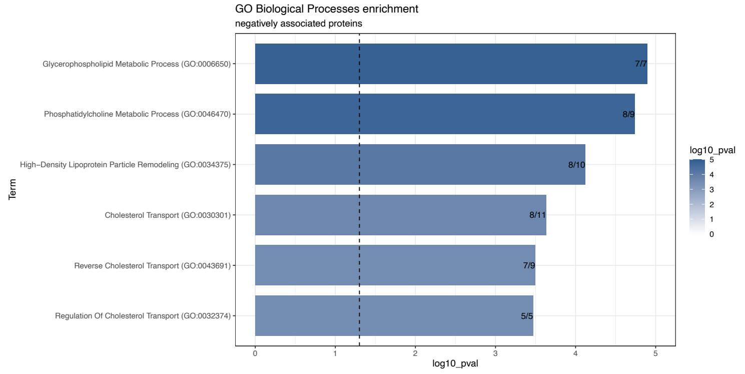


[D]

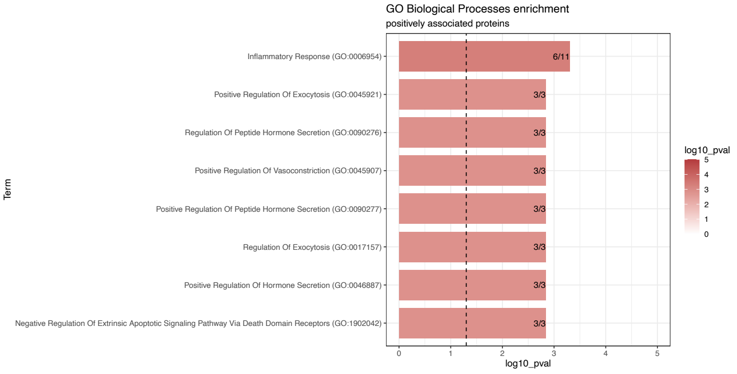


[E]

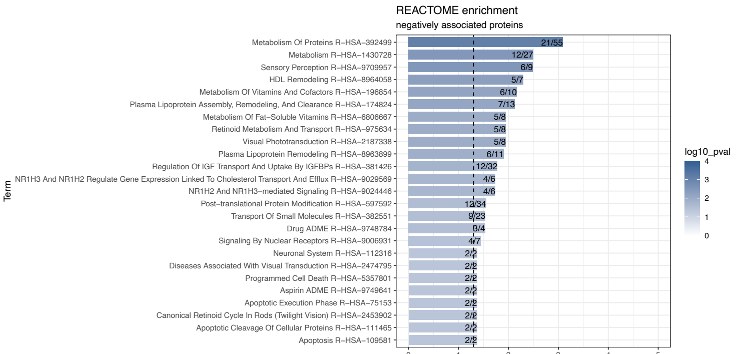


[F]

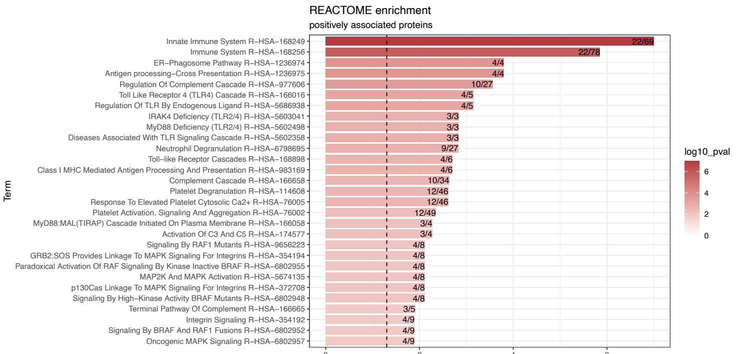

**Figure S8:** Covid19 related proteins show differences with MetaboHealth

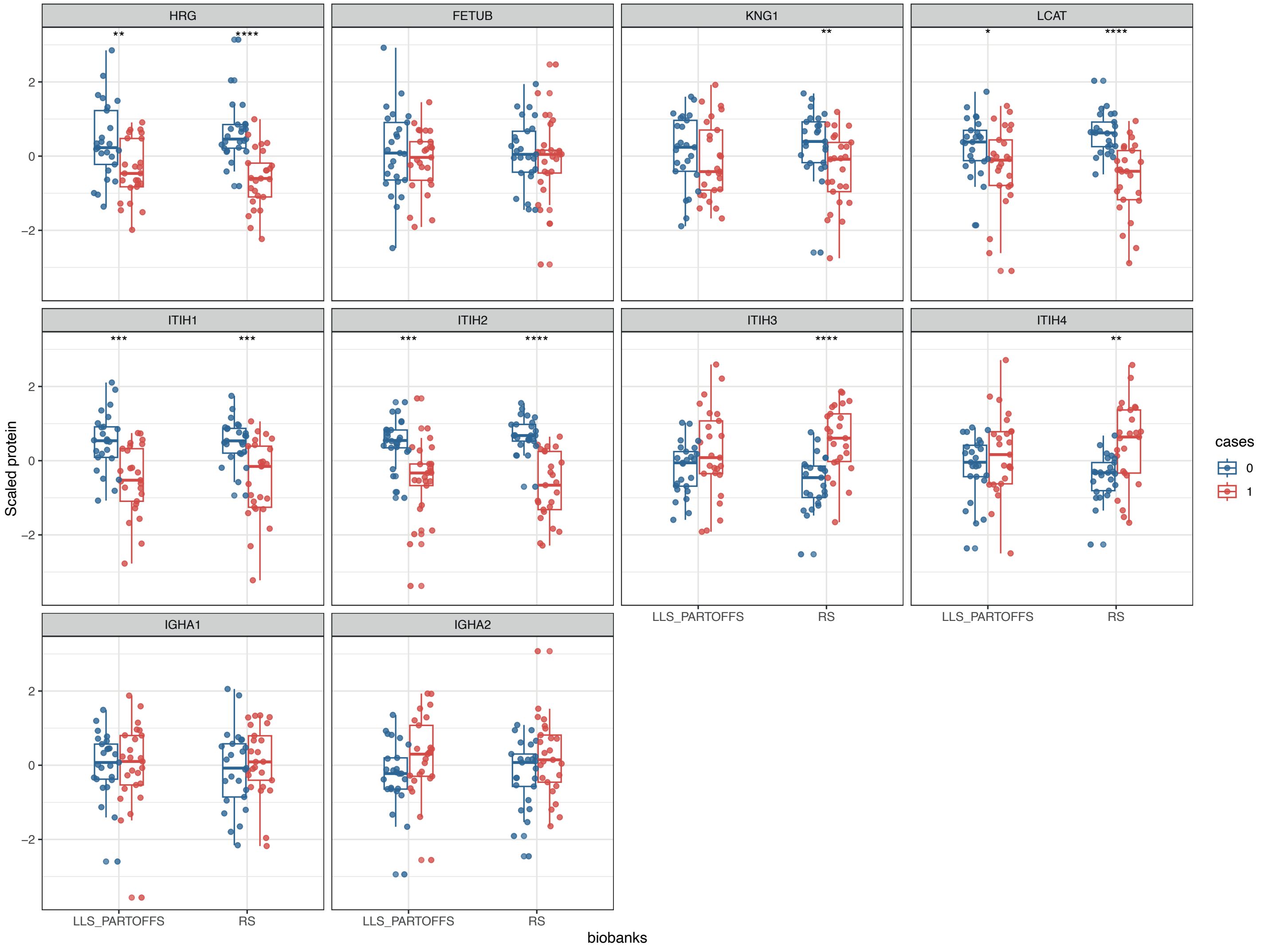

**Figure S9:** Preoprocessing in NTR

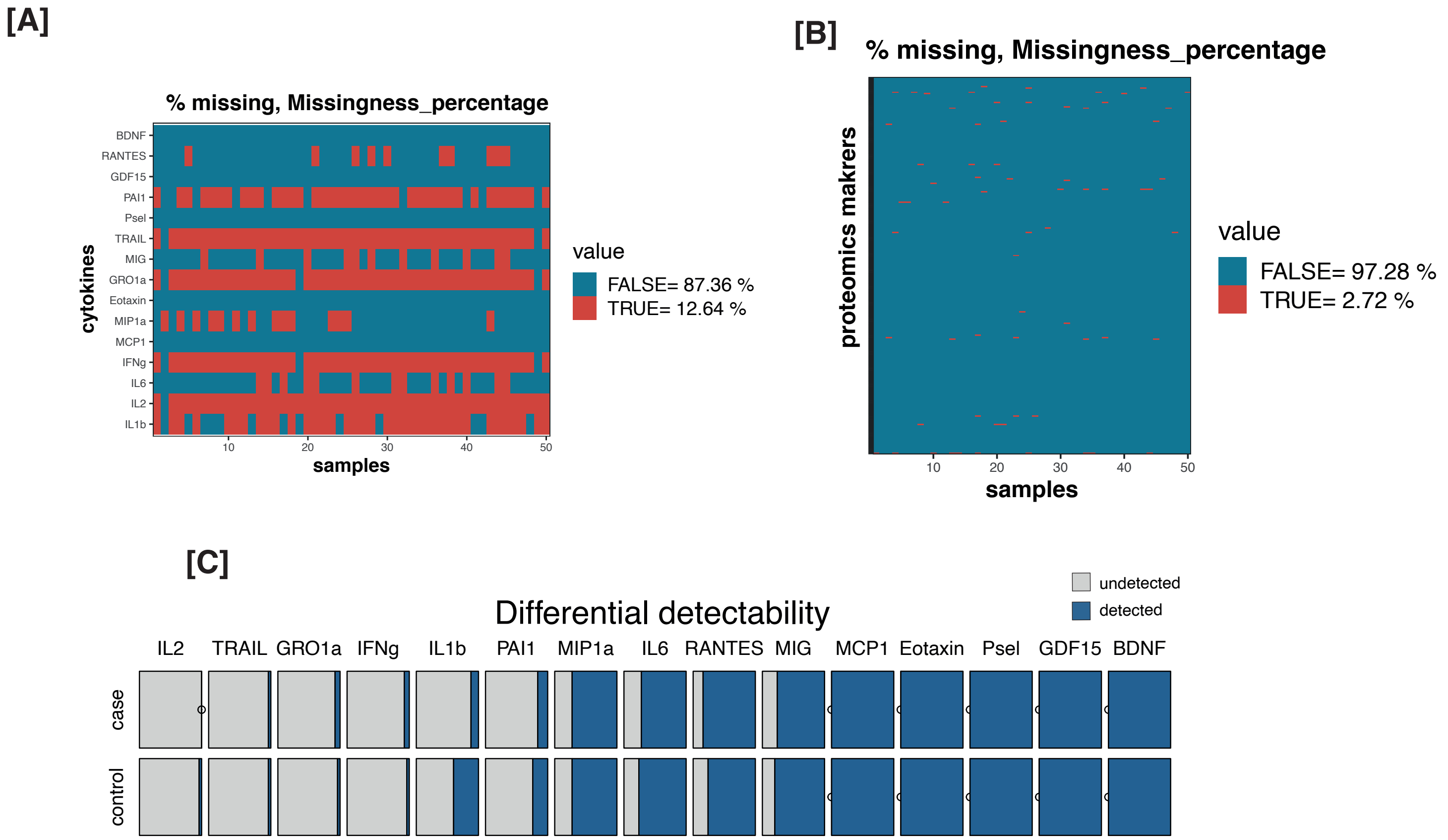

**Figure S10:** Associations in NTR

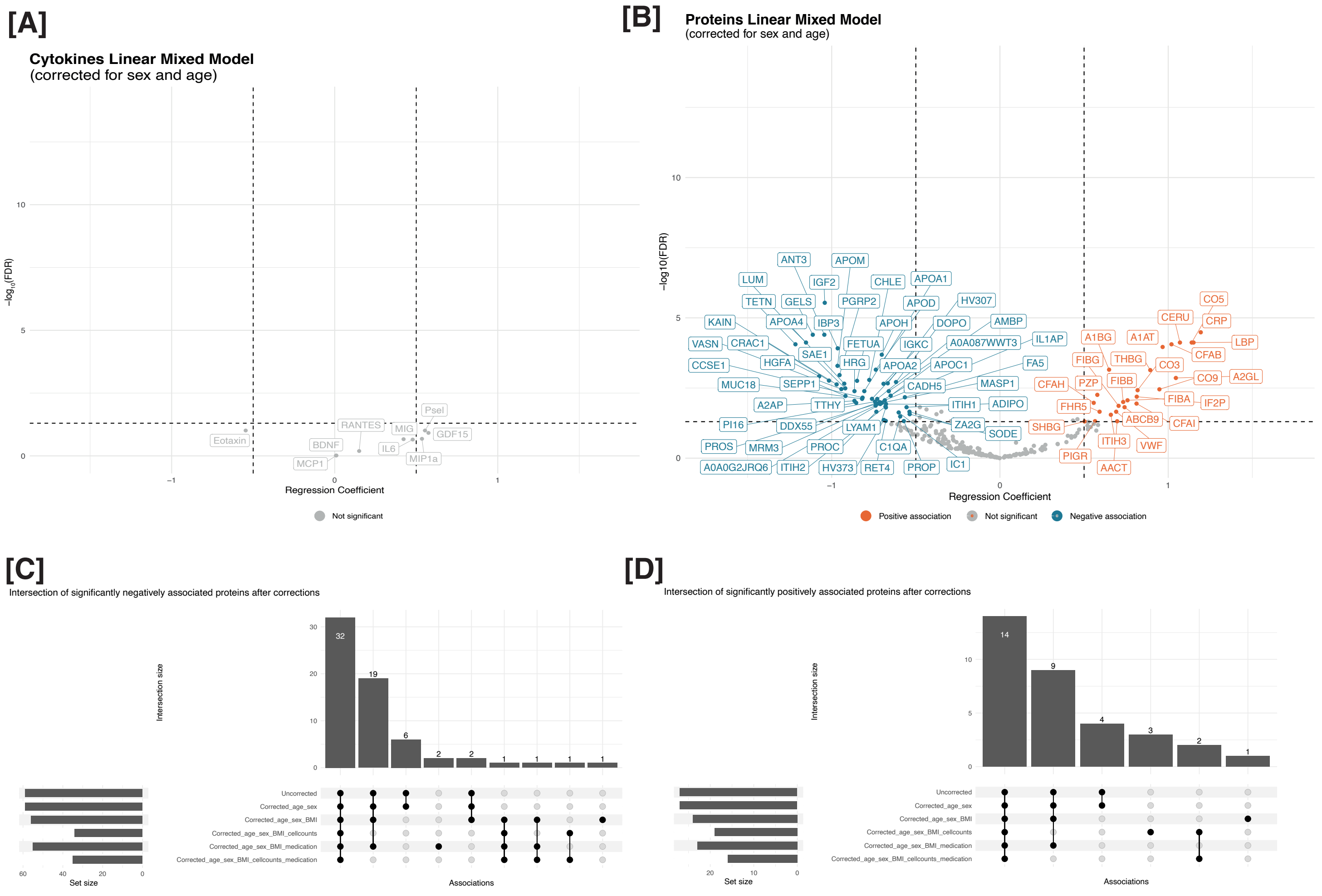

**Figure S11:** Complete heatmap profile of the significant proteins

**[A]**

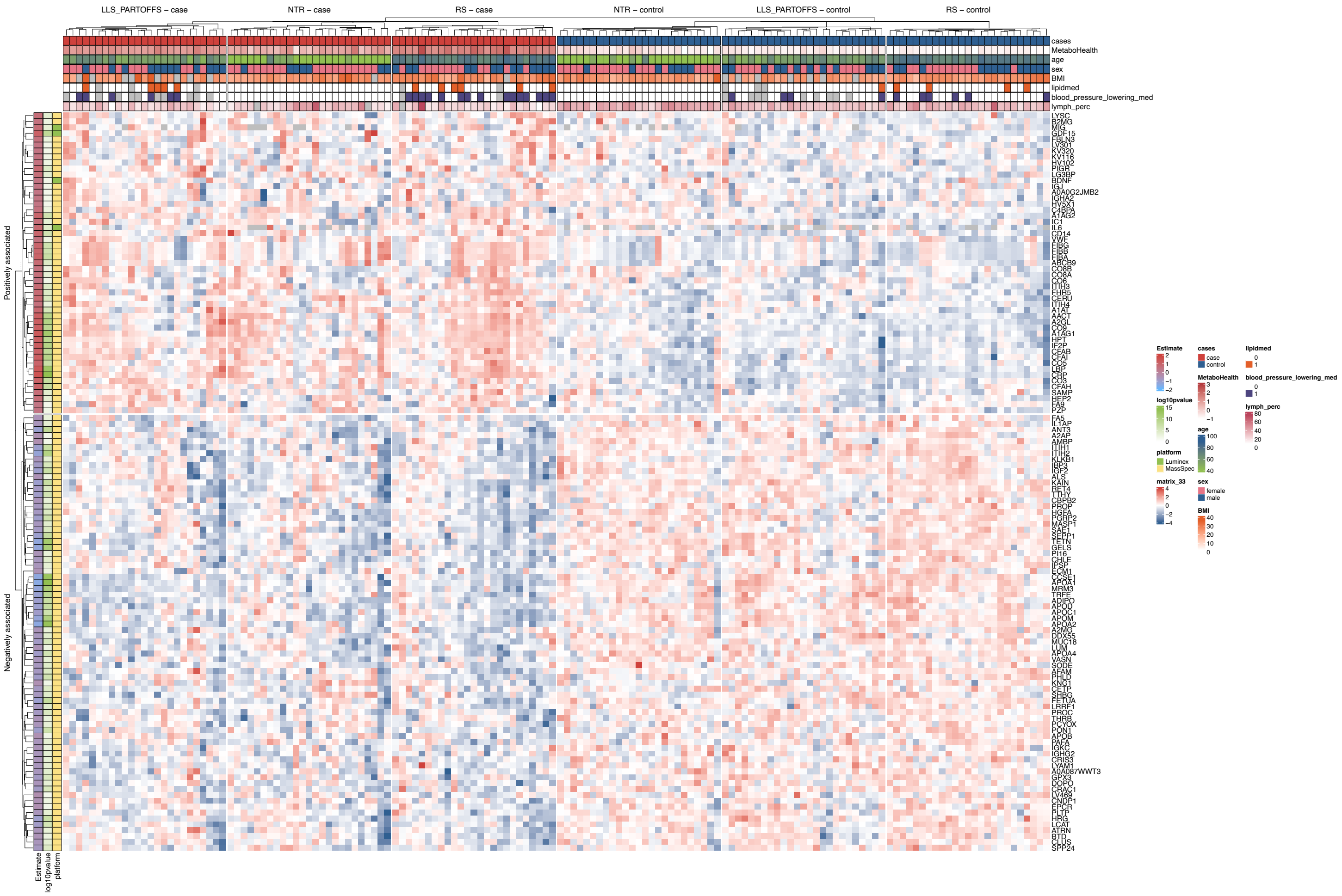
