## Supplementary Documents for "Extreme MetaboHealth scores in three cohort studies associate with plasma protein markers for inflammation and cholesterol transport"

**Supplementary Material**

Netherlands Twin Register ([NTR)	3](#_Toc163739779)

Netherlands Twin Register ([NTR)	3](#_Toc163739779)

Netherlands Twin Register ([NTR)	3](#_Toc163739779)

### Supplementary tables

Table S1: Phenotypic characteristics of the selected MetaboHealth Extremes.

| *Summary statistics LLS-PAROFFS*  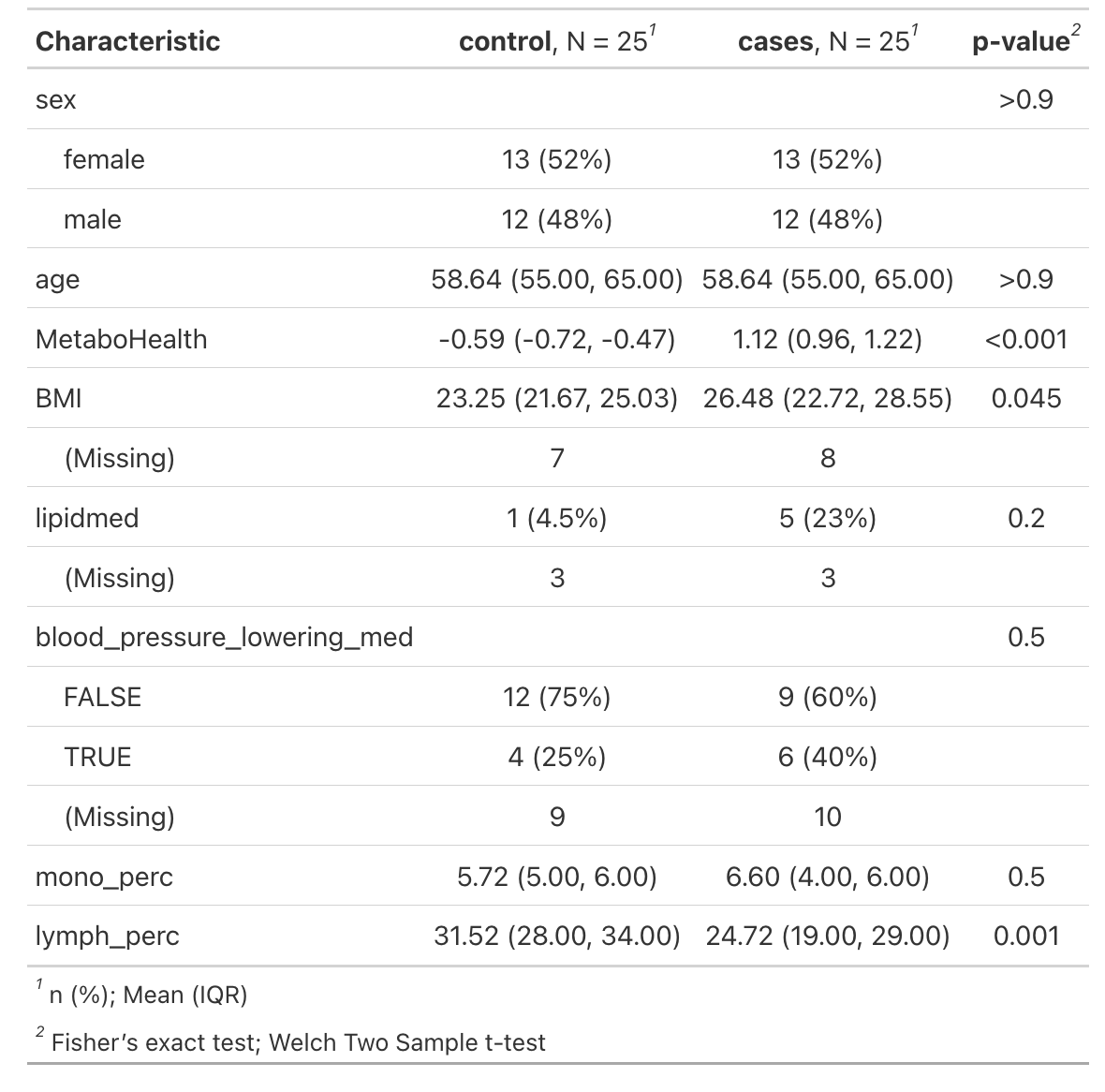 | *Summary statistics RS*  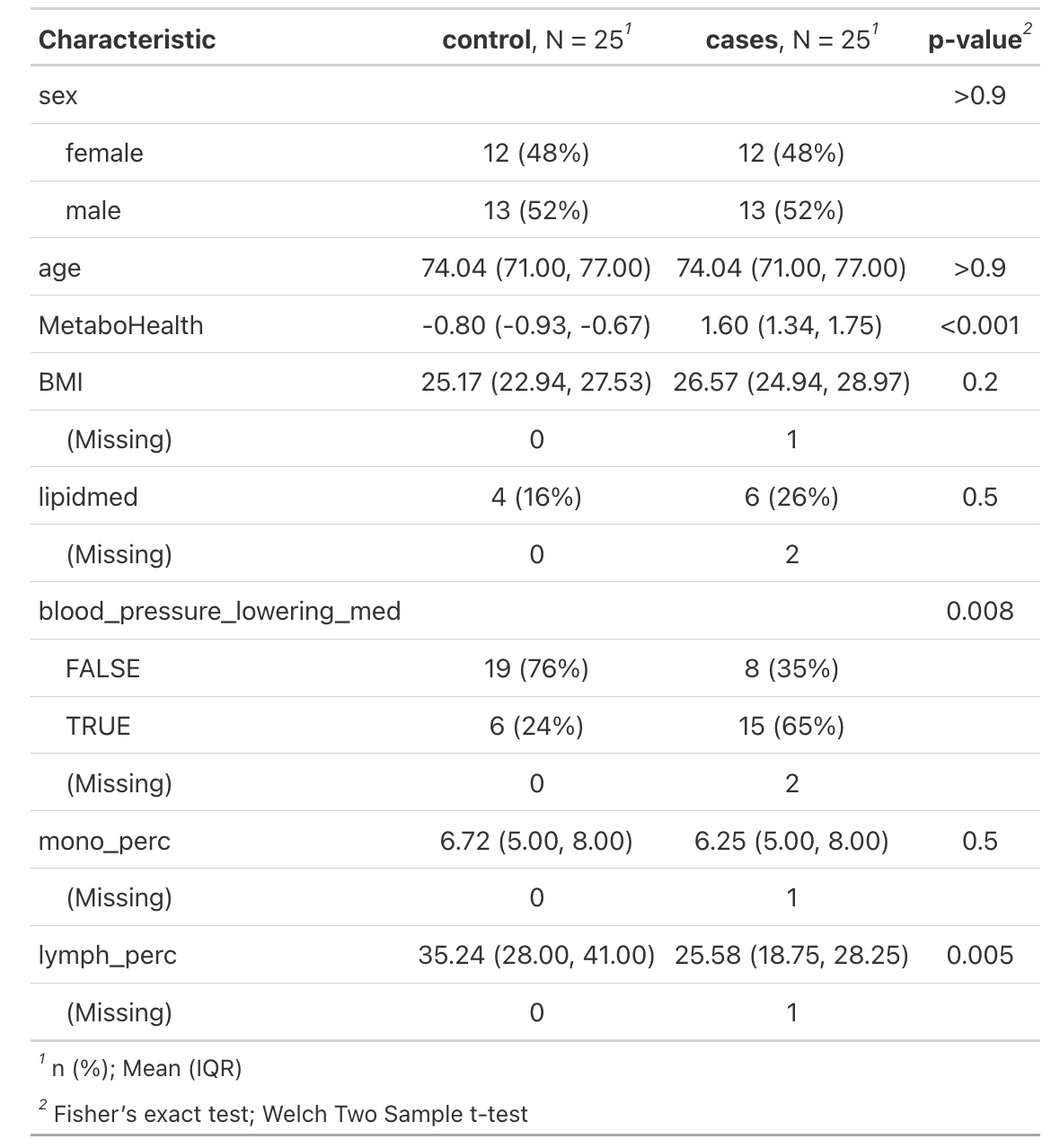 | *Summary statistics NTR*  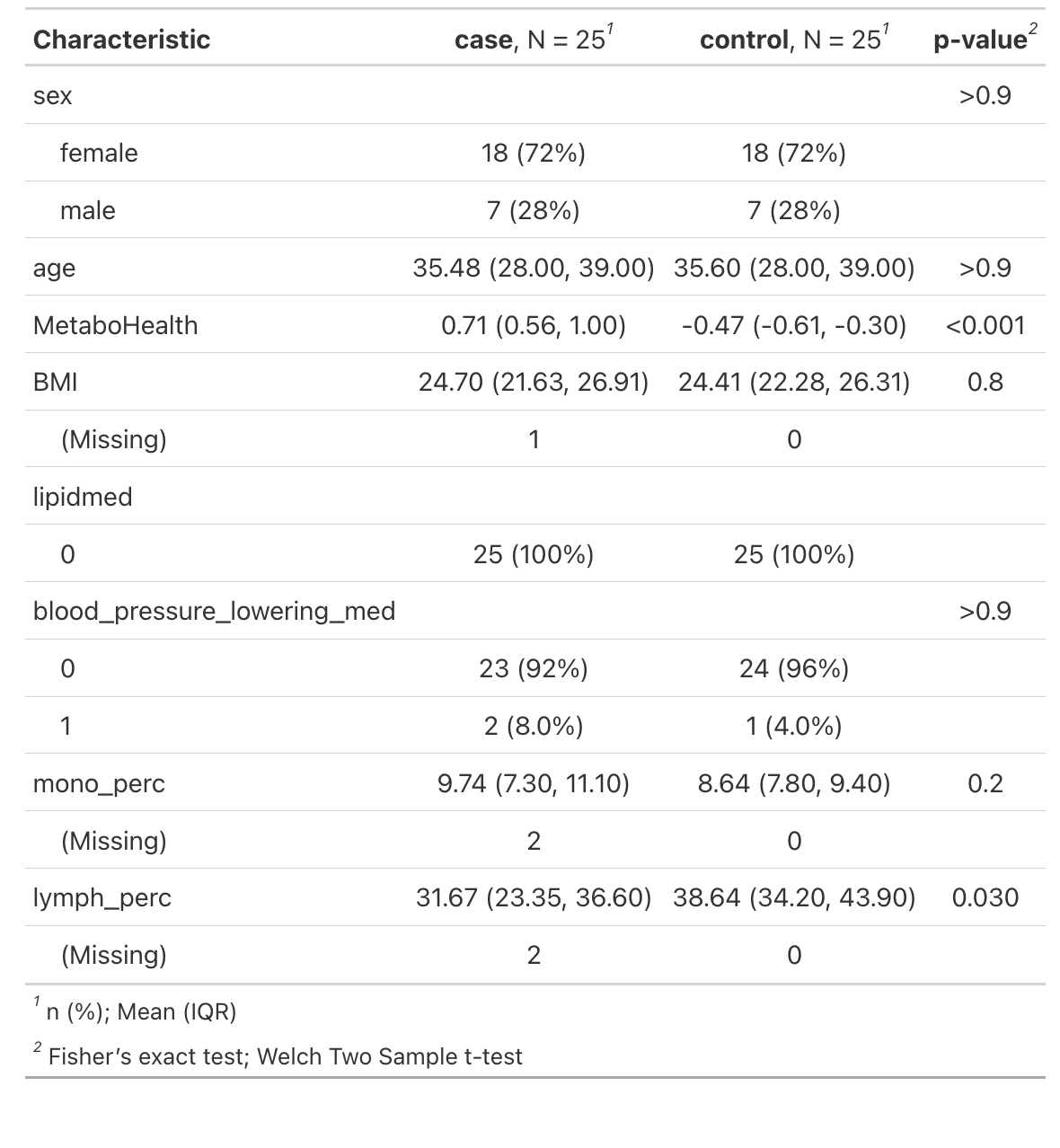 |
| --- | --- | --- |

### Consortium

#### Cohort Description

##### Leiden Longevity Study (LLS)

The Leiden Longevity Study (LLS) consists of 421 long-lived families of European descent. Families were included if at least two long-lived siblings were alive and fulfilled the age criterion of 89 years or older for males and 91 years or older for females, representing <0·5% of the Dutch population in 2001 [1]. In total, 944 long-lived proband siblings (mean age = 94 years, range = 89-104), 1671 offspring (mean age = 61 years, range = 39-81) and 744 spouses thereof (mean age = 60 years, range = 36-79) were included. Registry-based follow-up until the 27^th^ of October 2016 was available for all participants. Metabolites were successfully quantified in 843 nonagenarians [LLS_SIBS], 1157 of their offspring and 684 controls (LLS_PAROFFS) using non-fasted EDTA plasma samples.

1. M. Schoenmaker et al. Evidence of genetic enrichment for exceptional survival using a family approach: the Leiden Longevity Study. Eur J Hum Genet. 2006 Jan;14(1):79-84.

##### Rotterdam Study (RS)

The Rotterdam Study is a prospective, population-based cohort study among individuals living in the well-defined Ommoord district in the city of Rotterdam in The Netherlands [1]. The aim of the study is to determine the occurrence of cardiovascular, neurological, ophthalmic, endocrine, hepatic, respiratory, locomotor, dermatological, otolaryngological, and psychiatric diseases in elderly people. The cohort was initially defined in 1990 among approximately 7,983 persons, aged 55 years and older, who underwent a home interview and extensive physical examination at the baseline and during follow-up visits every 3-4 years (RS-I)[1]. Cohort was extended in 2000/2001 (RS-II, 3,011 individuals aged 55 years and older) and 2006/2008 (RS-III, 3,932 subjects, aged 45 and older). As of 2008, Rotterdam Study comprised 14,926 subjects. Written informed consent was obtained from all participants and the Medical Ethics Committee of the Erasmus Medical Center, Rotterdam, approved the study. Metabolomics measurements were quantified in fasted EDTA plasma samples using Nightingale Health platform. Metabolomics measurements were available for 2,986 participants from RS-I, 591 participants from RS-II (n=591) and 1,787 participants from RS-III.

1. Ikram, M. Arfan, et al. "The Rotterdam Study: 2018 update on objectives, design and main results." *European Journal of Epidemiology* 32.9 (2017): 807-850.

##### NTR

Since 1987, the Netherlands Twin Register is collecting (longitudinal) data in young and adult twins and their families [1,2]. The rich phenotypic longitudinal information that has been collected extends from lifestyle, exposures, personality and demographics information to mental and somatic health. In subgroups information on autonomic and central nervous system function, biomarkers and gene expression, epigenetics and genotyping is available. A 2015 estimate is that, since initiating the NTR, ~25% of all twins and multiples in the Netherlands participated in NTR research projects. Longitudinal information for over 200,000 participants (twins, multiples and family members) was collected over multiple NTR research projects. Data collection is ongoing. A pdf of nearly all published papers may be found at the NTR website.

As part of a Netherlands Twin Register (NTR) biobank project (BB1), 9,530 participants from 3,477 families were visited at home between January 2004 and July 2008 for collection of blood samples [3]. A second project (BB2) collected blood samples in 517 subjects from January 2011 to December 2011, including 210 MZ twin pairs and 64 twin-spouse pairs [4]. Visits were scheduled between 7:00 and 10:00 am and fertile women were bled on day 2-4 of the menstrual cycle, or in their pill-free week. Body composition was measured and information about physical health and lifestyle (e.g. smoking and drinking behavior, exercise, medication use) was obtained. For more detailed information about the methodology of the NTR Biobank study, see [3]. The NTR studies were approved by the Central Ethics Committee on Research involving human subjects of the VUMC, Amsterdam, an Institutional Review Board certified by the US Office of Human Research Protections (IRB number IRB-2991 under Federal wide Assurance-3703; IRB/institute codes, NTR 03-180). All subjects provided written informed consent. Subject were selected who were part of NTR biobank and who in general had rich phenotyping data available.

**website:** [www.tweelingenregister.org/](http://www.tweelingenregister.org/)

1. Van Beijsterveldt CE, Groen-Blokhuis M, Hottenga JJ, Franić S, Hudziak JJ, Lamb D, Huppertz C, de Zeeuw E, Nivard M, Schutte N, Swagerman S, Glasner T, van Fulpen M, Brouwer C, Stroet T, Nowotny D, Ehli EA, Davies GE, Scheet P, Orlebeke JF, Kan KJ, Smit D, Dolan CV, Middeldorp CM, de Geus EJ, Bartels M, Boomsma DI. The Young Netherlands Twin Register (YNTR): longitudinal twin and family studies in over 70,000 children. *Twin Res Hum Genet.* **2013** Feb; 16(1): 252-267. [doi: 10.1017/thg.2012.118](https://doi.org/10.1017/thg.2012.118). PMID: 23186620

2. Willemsen G, Vink JM, Abdellaoui A, den Braber A, van Beek JH, Draisma HH, van Dongen J, van 't Ent D, Geels LM, van Lien R, Ligthart L, Kattenberg M, Mbarek H, de Moor MH, Neijts M, Pool R, Stroo N, Kluft C, Suchiman HE, Slagboom PE, de Geus EJ, Boomsma DI.The Adult Netherlands Twin Register: twenty-five years of survey and biological data collection.*Twin Res Hum Genet.* **2013** Feb; 16(1): 271-281. [doi: 10.1017/thg.2012.140](https://doi.org/10.1017/thg.2012.140). PMID: 23298648. PMCID: PMC3739974.

3. Willemsen G, de Geus EJ, Bartels M, van Beijsterveldt CE, Brooks AI, Estourgie-van Burk GF, Fugman DA, Hoekstra C, Hottenga JJ, Kluft K, Meijer P, Montgomery GW, Rizzu P, Sondervan D, Smit AB, Spijker S, Suchiman HE, Tischfield JA, Lehner T, Slagboom PE, Boomsma DI. The Netherlands Twin Register biobank: a resource for genetic epidemiological studies. *Twin Res Hum Genet.* **2010** Jun; 13(3): 231-245. [doi: 10.1375/twin.13.3.231](https://doi.org/10.1375/twin.13.3.231). PMID: 20477721.

4. Sirota M, Willemsen G, Sundar P, Pitts SJ, Potluri S, Prifti E, Kennedy S, Ehrlich SD, Neuteboom J, Kluft C, Malone KE, Cox DR, de Geus EJ, Boomsma DI. Effect of genome and environment on metabolic and inflammatory profiles. *PLoS One.* **2015** Apr 8; 10(4): e012089. [doi: 10.1371/journal.pone.0120898](https://doi.org/10.1371/journal.pone.0120898). PMID: 25853885. PMCID: PMC4390246.

5 van Dongen J, Nivard MG, Willemsen G, Hottenga JJ, Helmer Q, Dolan CV, Ehli EA, Davies GE, van Iterson M, Breeze CE, Beck S, Bios Consortium, Suchiman HE, Jansen R, van Meurs JB, Heijmans BT, Slagboom PE, Boomsma DI. Genetic and environmental influences interact with age and sex in shaping the human methylome. Nature Communication. 2016 Apr 7; 7(1): 11115. doi: 10.1038/ncomms11115 (2016). PMID: 27051996. PMCID: PMC4820861.

#### Acknowledgements

##### Leiden Longevity Study (LLS)

The LLS has received funding from the European Union's Seventh Framework Programme (FP7/2007-2011) under grant agreement n° 259679. This study was supported by a grant from the Innovation-Oriented Research Program on Genomics (SenterNovem IGE05007), the Centre for Medical Systems Biology, and the Netherlands Consortium for Healthy Ageing (grants 05040202 and 050-060-810), all in the framework of the Netherlands Genomics Initiative, Netherlands Organization for Scientific Research (NWO), Unilever Colworth, and by BBMRI-NL, a Research Infrastructure financed by the Dutch government (NWO 184.021.007 and 184.033.111).

##### Rotterdam Study (RS)

The Rotterdam Study is supported by the Erasmus MC University Medical Center and Erasmus University Rotterdam; The Netherlands Organisation for Scientific Research (NWO); The Netherlands Organisation for Health Research and Development (ZonMw); the Research Institute for Diseases in the Elderly (RIDE); The Netherlands Genomics Initiative (NGI); the Ministry of Education, Culture and Science; the Ministry of Health, Welfare and Sports; the European Commission (DG XII); and the Municipality of Rotterdam. The authors are grateful to the study participants, the staff from the Rotterdam Study and the participating general practitioners and pharmacists. Metabolomics measurements were funded by Biobanking and Biomolecular Resources Research Infrastructure (BBMRI)–NL (184.021.007) and the JNPD under the project PERADES (grant number 733051021, Defining Genetic, Polygenic and Environmental Risk for Alzheimer’s Disease using multiple powerful cohorts, focused Epigenetics and Stem cell metabolomics).

##### Nethelands Twin Register (NTR)

Funding was obtained from the Netherlands Organization for Scientific Research (NWO) and MagW/ZonMW grants 904-61-090, 985-10-002, 904-61-193,480-04-004, 400-05-717, Addiction-31160008, Middelgroot-911-09-032, Spinozapremie 56-464-14192, Biobanking and Biomolecular Resources Research Infrastructure (BBMRI –NL, 184.021.007).; the European Community's Seventh Framework Program (FP7/2007-2013), ENGAGE (HEALTH-F4-2007-201413); the European Science Council (ERC Advanced, 230374), Rutgers University Cell and DNA Repository (NIMH U24 MH068457-06), the Avera Institute, Sioux Falls, South Dakota (USA) and the National Institutes of Health (NIH, R01D0042157-01A, MH081802, Grand Opportunity grants 1RC2 MH089951). We gratefully acknowledge grant NWO 480-15-001/674: Netherlands Twin Registry Repository: researching the interplay between genome and environment.

#### Ethics statements

##### Leiden Longevity Study (LLS)

The Leiden Longevity Study protocol was approved by the Medical Ethical Committee of the Leiden University Medical Center before the start of the study (P01.113). In accordance with the Declaration of Helsinki, the Leiden Longevity Study obtained informed consent from all participants prior to their entering the study.

##### Rotterdam Study

*The Rotterdam S*tudy protocol was approved by the Medical Ethics Committee of the Erasmus MC Rotterdam, the Nethrlands. (*MEC 02.1015*) and by the Dutch Ministry of Health, Welfare, and Sport (Population Screening Act WBO, license number 1071272-159521-PG). In accordance with the Declaration of Helsinki, the *Rotterdam Study* obtained written informed consent from all participants prior to their entering the study.

##### VUNTR

The Netherlands twin Register study protocol was approved by the Central Ethics Committee on Research Involving Human Subjects of the VU University Medical Centre, Amsterdam, an Institutional Review Board certified by the U.S. Office of Human Research Protections (IRB number IRB00002991 under Federal-wide Assurance- FWA00017598. In accordance with the Declaration of Helsinki, the Netherlands Twin Register obtained informed consent from all participants prior to their entering the study.
